# Early and ongoing importations of SARS-CoV-2 in Canada

**DOI:** 10.1101/2021.04.09.21255131

**Authors:** Angela McLaughlin, Vincent Montoya, Rachel L. Miller, Gideon J. Mordecai, Michael Worobey, Art F. Y. Poon, Jeffrey B. Joy

## Abstract

Tracking the emergence and spread of SARS-CoV-2 is critical to inform public health interventions. Phylodynamic analyses have quantified SARS-CoV-2 migration on global and local scales^1–5^, yet they have not been applied to determine transmission dynamics in Canada. We quantified SARS-CoV-2 migration into, within, and out of Canada in the context of COVID-19 travel restrictions. To minimize sampling bias, global sequences were subsampled with probabilities corrected for their countries’ monthly contribution to global new diagnoses. A time-scaled maximum likelihood tree was used to estimate most likely ancestral geographic locations (country or Canadian province), enabling identification of sublineages, defined as introduction events into Canada resulting in domestic transmission. Of 402 Canadian sublineages identified, the majority likely originated from the USA (54%), followed by Russia (7%), India (6%), Italy (6%), and the UK (5%). International introductions were mostly into Ontario (39%) and Quebec (38%). Among Pango lineages^6^, B.1 was imported at least 191 separate times from 11 different countries. Introduction rates peaked in late March then diminished but were not eliminated following national interventions including restrictions on non-essential travel. We further identified 1,380 singleton importations, international importations that did not result in further sampled transmission, whereby representation of lineages and location were comparable to sublineages. Although proportion of international transmission decreased over time, this coincided with exponential growth of within-province transmission – in fact, total number of sampled transmission events from international or interprovincial sources increased from winter 2020 into spring 2020 in many provinces. Ontario, Quebec, and British Columbia acted as sources of transmission more than recipients, within the caveat of higher sequence representation. We present strong evidence that international introductions and interprovincial transmission of SARS-CoV-2 contributed to the Canadian COVID-19 burden throughout 2020, despite initial reductions mediated by travel restrictions in 2020. More stringent border controls and quarantine measures may have curtailed introductions of SARS-CoV-2 into Canada and may still be warranted.

**Significance Statement:** By analyzing SARS-CoV-2 genomes from Canada in the context of the global pandemic, we illuminate the extent to which the COVID-19 burden in Canada was perpetuated by ongoing international importations and interprovincial transmission throughout 2020. Although travel restrictions enacted in March 2020 reduced the importation rate and proportion of transmission from abroad across all Canadian provinces, SARS-CoV-2 introductions from the USA, India, Russia, and other nations were detectable through the summer and fall of 2020.

## Introduction

The SARS-CoV-2 pandemic has highlighted the importance of genomic epidemiology in deciphering the origin and spread of emerging pathogens across local, national, and global scales to aid in directing responses. These analyses are entirely dependent upon the timely generation, assembly, and sharing of publicly available genetic sequences and associated metadata through the Global Initiative on Sharing All Influenza Data (GISAID) platform. The global effort to sequence and share SARS-CoV-2 genomes has been unprecedented, such that by 11 February 2021, just over a year since the first whole genome sequence was shared^7,8^, there were 495,159 SARS-CoV-2 sequences publicly available on GISAID representing 108,301,802 confirmed COVID-19 diagnoses^9^. Studying evolutionary relationships between SARS-CoV-2 samples over time and space using molecular phylogenetics illuminates underlying epidemic dynamics, such as the relative contributions of international and domestic transmission, and in doing so can be highly informative in evaluating whether particular non-pharmaceutical interventions have been effective in curbing importations from abroad, as well as transmission within jurisdictions.

Although SARS-CoV-2 exhibits a relatively slow substitution rate on a per site basis compared to other RNA viruses, its evolutionary rate per genome is comparable or high due to its large genome of 29 kb compared to the average of 9 kb^10^, as well as its high transmission rate. Thus ample divergence has accumulated to distinguish globally sampled viruses into groups defined by common mutations and recent ancestry; the most commonly applied nomenclature include clades as defined by Nextstrain^19,20^ and phylogenetic Pango lineages^6^. Tracking phylogenetic lineages’ spatiotemporal movement is useful to better understand the relative contributions of importations and local transmission. The importance of partitioning sequences into a dynamic and accurate nomenclature system is critical because it allows a common framework for genomic surveillance and for discussion of emerging variants of concern (VOC)^6,11^.

Tracking the dynamics and genomic characteristics of VOC – notably B.1.351, B.1.1.7, and P.1 - has become a critical global concern in light of their potential to hamper the effectiveness of vaccines and non-pharmaceutical interventions. In early August 2020, the B.1.351 variant, also known as 501Y.V2, was first detected in South Africa^12^; this was quickly followed by the identification of the B.1.1.7 variant, which was first sampled in Southern England in September 2020 and rapidly rose in frequency^11^. Shortly thereafter, the P.1 variant (alias: B.1.1.28.1), emerged in Manaus, Brazil in early December 2020^13^. As the B.1.351 and P.1 lineages harbour E484K, K417T, and N501Y, it seems likely that they display elevated capacity for immune evasion^13^. The 69-70 deletion in spike, which co-occurs with N501Y in B.1.1.7, has also been implicated in immune escape^14,15^. The rapid global emergence and spread of VOC and the risk they pose towards the effectiveness of non-pharmaceutical interventions and vaccines emphasizes the importance of identifying and tracking novel variants and their introduction into new geographical regions.

Relatively early in the COVID-19 epidemic, on 25 January 2020, the first case in Canada was detected in a traveller from Wuhan to Toronto, where it was suspected that onward transmission was limited to his wife^16^(Fig. S1). It was not until 20 February that Canada reported its first case related to travel from outside mainland China. In the province of British Columbia, sequences from within the lineages A and A.1, which diverged from the B lineages early in the pandemic, were identified through March and April, alongside international events in Vancouver, British Columbia, namely an international dentistry conference 5-7 March where at least 42 people were infected^17^ and the World Rugby 7s on 7–9 March. Another notable Canadian superspreader event simultaneously occurred in Edmonton, Alberta at a curling bonspiel (tournament) on 11 March attended by doctors from across Western Canada^18^, where 45 out of 70 participants were infected. In the subsequent week, the stringency of Canadian interventions increased rapidly (Fig. S1). On 14 March, a travel advisory warning against all non-essential travel outside Canada was issued; on 18 March, travel restrictions on the entry of all foreign nationals (except from the United States) were announced; 21 March travel restrictions were extended to the USA; and 24 March, the mandatory 14-day self-isolation for those returning from international travel was announced. By 9 June, travel restrictions on foreign nationals with immediate family in Canada relaxed^19^. Studies of the genomic epidemiology of SARS-CoV-2 in Canada have been heretofore been limited to a single study focusing exclusively on the early epidemic in Quebec^20^. Murall *et al*. conservatively estimated more than 200 independent events into Quebec by late March, suggesting that international introductions in were largely attributable to the province’s “spring break”, which occurred 2-3 weeks earlier than the rest of Canada^20^. Similar analyses have not yet been applied at a national scale, and the relative contributions of international introductions and interprovincial transmission in sustaining the COVID pandemic in Canada are not well-understood, despite their potential importance in responding to future pandemics or resurgences of SARS-CoV-2. Despite some common interventions, provincial and territorial cumulative incidences (Fig. S2) and response stringencies^21^ have drastically differed; thus, we expect that viral evolutionary dynamics should also vary.

Phylogeographic methods to infer ancestral states of sampled viruses have been applied widely to quantify the timing and geographic origins of SARS-CoV-2 introductions into the United Kingdom (UK)^5^, United States of America (USA)^1–3,22^, Brazil^23,24^, New Zealand (NZ)^4,25^, and Europe^1,26^, among others. Drastically different epidemic dynamics have been elucidated globally. For example, in the UK, with the highest sequences per case of any country^27^, du Plessis *et al*. conservatively estimated 1,179 independent introductions resulting in 2 or more sampled descendant cases^5^. Importantly, their analyses revealed that detection lag – the amount of time between the time of the most recent common ancestor (tMRCA) and the first sample collection date for a given UK transmission lineage – decreased over time as more genomes were generated, suggesting that efficiency in detecting and responding to importations improved over time. An analysis of early epidemic dynamics in the state of Louisiana, USA, revealed starkly contrasting importation dynamics, wherein the first wave of SARS-CoV-2 was almost entirely attributable to a single domestic introduction. This introduction likely originated from Texas, which occurred several weeks prior to the annual Mardi Gras festival, during which superspreader events likely gave rise to a large number of cases within Louisiana and neighbouring Southern states^22^. Furthermore, in NZ, where border closures and stringent lockdown measures were enacted early in the pandemic^4^, reductions in international introductions and in reproductive numbers following lockdown were proportional to the reduction in mobility nationwide^4^. Illustrating the challenges of incorporating phylogenetic methods into public health responses, their analyses also revealed instances where individuals who were previously considered connected through contact tracing were not phylogenetically proximate or linked through a recent transmission event, and conversely, individuals’ viruses being highly similar despite no known contact. Geoghegan *et al*. further corroborated the effectiveness of the public health response in NZ, as they estimated that of 227 inferred introductions to NZ up to July 1 2020, 24% led to one downstream case and only 19% resulted in more than one downstream case^4^. Similar analyses are yet to be conducted for Canada.

Here we estimated the timing, origins, and destinations of SARS-CoV-2 introductions in Canada throughout 2020 for both singleton importations with no further sampled transmission and Canadian sublineages, which are introductions that ignited sampled domestic outbreaks. We applied a maximum likelihood approach to test the hypotheses that international importations of SARS-CoV-2 into Canada were reduced following the implementation of Canada-wide travel restrictions, as well as whether international importations contributed to ongoing transmission throughout the spring, summer, and fall of 2020. We further tested the hypotheses that over time testing and tracing may have improved such that there were fewer importations resulting in onward domestic transmission, smaller sublineages, and smaller delays in identifying new sublineages. These analyses contribute to a finer resolution understanding of how international importations acted as sparks that ignited ongoing SARS-CoV-2 outbreaks throughout Canada in 2020.

## Methods

### Timeline of COVID-19 in Canada

The dates of national-level COVID-19 interventions were obtained from the Canadian Institute for Health Information^19^ and the key events were obtained from a summary published by the National Post^16^. The Stringency Index for Canada was obtained from the Oxford COVID-19 Government Response Tracker^28^ (Fig. S1).

### Data cleaning

A total of 495,159 SARS-CoV-2 sequences were downloaded from GISAID with associated metadata, of which 9,862 were sampled in Canada, on 11 February 2021 (www.gisaid.org; contributing labs in Supplementary Appendix). Sequences were aligned pairwise to the Wuhan-Hu-1 reference sequence (Genbank Accession ID: MN908947.3) using the viralMSA wrapper tool invoking minimap2^29,30^. Subsequently, sequences were removed from the analysis if they were listed in the Nextstrain exclusion list on the day of data download^31,32^. This list identifies duplicate depositions (i.e., multiple samples from the same individual) or samples that contained known sequencing issues (n=788), contained >20% Ns (n=198) or >10% gaps (n=21,133, including 44 that also had high Ns), were sampled from a non-human host (n=925), were environmental samples (n=123), were likely to contain sequencing errors based on previous temporal analyses (n=2)^33^, or had incomplete sample collection dates (n=5,795). Canadian sequences with incomplete collection dates (n=2,168) were retained, however 82 of them were removed as they also lacked the month of collection, and the missing dates were inferred using LSD2 within IQ-TREE 2.1.2^35,36^ while converting the tree with branch lengths scaled in substitutions/site into a time-scaled tree under a relaxed molecular clock (see phylogenetic inference below). More recently, updated full sample collection dates from Japan (n=15,997) and the USA (n=1) were added to the metadata prior to excluding incomplete dates.

There were 466,025 clean sequences remaining, of which 9,657 were Canadian. Problematic sites (v4.3) identified by de Maio *et al*.^34^ were masked in the alignment. These sites include the untranslated ends of the alignment, which have low coverage and a high rate of sequencing or mapping errors, highly homoplasic sites with low prevalence and/or low phylogenetic signal, homoplasic sites specific to particular locations or labs, and homoplasic sites with strong phylogenetic signal. Nextstrain clade and Pango lineage designations were made before masking sites.

### Subsampling sequences

To minimize the impact of sampling bias on our analyses and minimise computational effort, we randomly sampled sequences to adjust for countries’ proportional contribution to new global COVID-19 cases in each month. Using the coronavirus R package^9^, country-specific new diagnosis counts by day were aggregated by month. For each calendar month, the proportion of total new diagnoses in each country among all new global diagnoses (minus Canada) was applied as a sampling probability for sequences from that month (Fig. 1a). Sequences were sampled without replacement. We strove to equalize the number of sequences sampled monthly; however there were relatively few sequences available until March 2020. Therefore, all clean sequences in the preceding months were included and the remainder of sequences sampled were distributed among well-sampled months (Table S1). The number of clean sequences available by month for Canadian provinces is summarized in Table S2. We mapped provinces with available sequences and annotated their cumulative per capita incidence (Fig. S2) using population data and NAD83 shapefiles from the Statistics Canada 2016 Census^37,38^ and NAD83 shapefiles for the USA from the United States Census Bureau^39^. The number of cases by Canadian province over time was obtained from the Public Health Agency of Canada^40^.

**Fig. 1.**
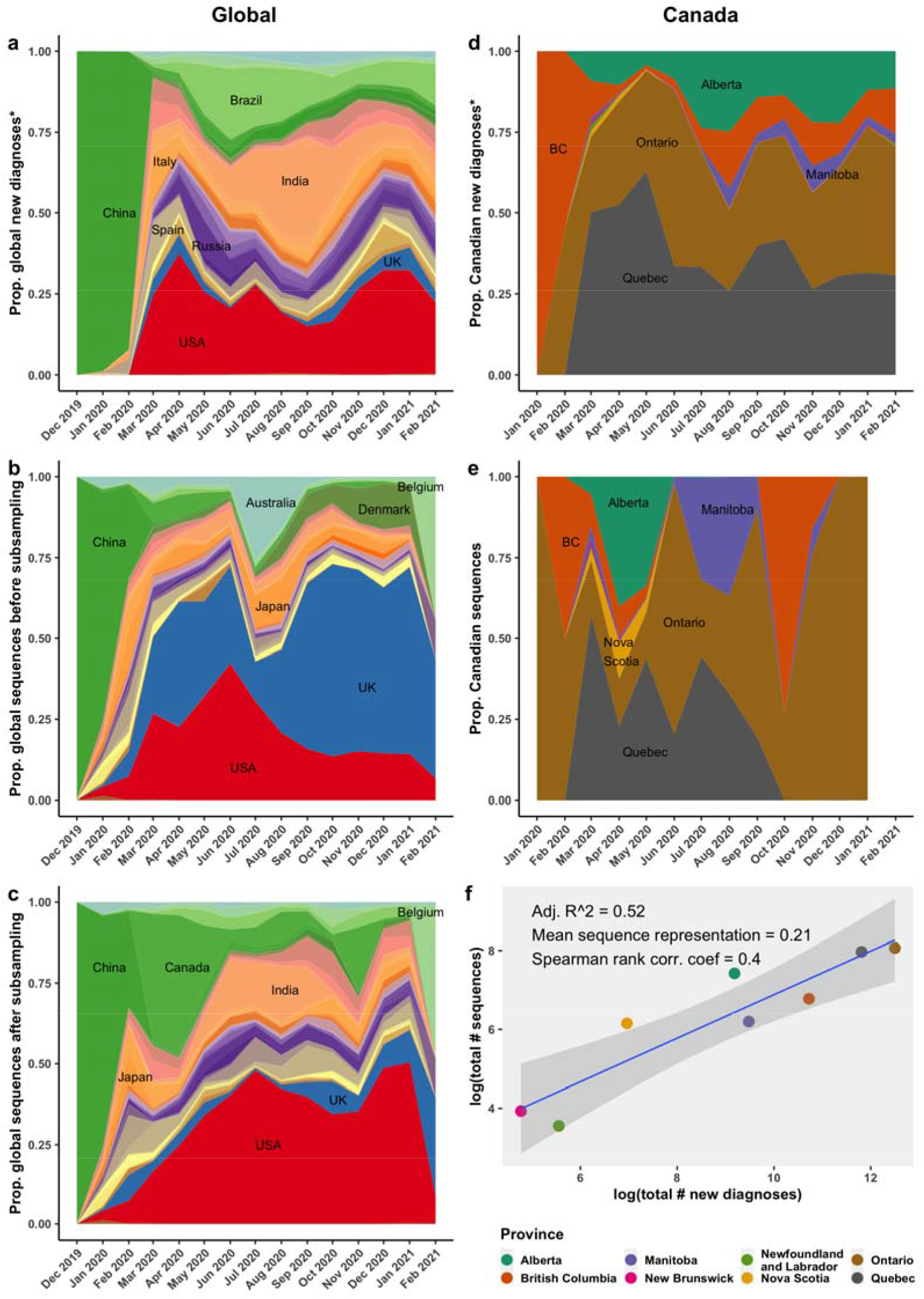
Canadian sequences were analyzed in the context of global sequences subsampled with probabilities reflecting countries’ monthly contributions to global new diagnoses. Countries’ a) monthly contributions to global new diagnoses (*among countries with sequences available), b) monthly proportional contributions to cleaned GISAID sequences before subsampling, and c) monthly contributions to sequences after subsampling. All Canadian clean sequences were kept for the analysis, causing a discrepancy between provinces’ d) monthly contributions to Canadian new diagnoses (*among provinces with sequences available) and e) monthly contributions to Canadian sequences. f) The overall relationship between total number of clean sequences and total number of confirmed diagnoses by province. Spearman rank correlation coefficient was calculated without log transformations.

With the aim of subsampling 50,000 sequences total for computational feasibility while focusing on Canadian dynamics, we sampled all 9,657 clean Canadian sequences and the remainder of 40,333 from the pool of global clean sequences (n=456,368). The probability of selecting any given global sequence was adjusted for its country’s contribution to monthly global new diagnoses, excluding Canada. To account for underrepresentation of sequences from Iran and Italy early in the epidemic despite their known importance in early transmission dynamics^1^, sequences from individuals who had travel history to Iran or Italy were recategorized as having been sampled in the country of travel if the sampling date predated June 2020 – this affected five Canadian sequences across February and March 2020. The relative contribution of geographies to the global sequence set before and after subsampling is summarized in Fig. 1b-c, while the overall sequence representation (# sequences / # confirmed diagnoses) for each geography before and after subsampling is shown in Fig. S3.

### Maximum likelihood phylogenetic inference

We used the subsampled alignment to infer an approximate maximum likelihood (ML) phylogeny using FastTree version 2.2.1^41^ with a generalized time-reversible substitution model with random starting tree seeds. The tree was rooted using R package ape^42^ on the earliest lineage A sequence (EPI_ISL_406801, sampled 05 January 2020) based on the presence of two nucleotides not found in the commonly used reference (Wuhan-Hu-1, sampled 26 December 2019), which are shared with the most proximate bat coronaviruses identified thus far, RaTG13 and RmYN02^6^. We then fit a slope of evolutionary distance (number of inferred substitutions between root and tips) over time using Tempest^43^ and excluded temporal outliers with residuals further than the mean residual plus or minus 3 standard deviations (n=385) (Fig. S4a). To further clean the data of temporal outliers, any tips with pendant edges (edge immediately preceding tip) with more than 12 mutations after masking were removed (n=328), under the justification that it would take about 6 months for 12 mutations to accumulate if we assume a strict molecular clock of 8e-4 substitutions/site/year (Fig. S4b). The molecular clock rate fit before and after removing temporal outliers is shown in Fig. S4c-d. The cleaned tree in units of substitutions/site was converted to a time-scaled tree using LSD2 invoked within IQ-TREE 2.1.2, specifying a lognormal relaxed clock with 0.2 standard deviations, as well as a generalized time-reversible substitution model with gamma rate variation, invariant sites, and three discrete rate categories (GTR+I+R3), which was identified as the best-fitting model for a global phylogeny^44^, Wuhan-Hu-1 as outgroup (used because of its earlier sample date than EPI_ISL_406801), and 100 bootstraps to calculate confidence intervals on inferred dates^35,36^(Fig. S10). A total of 2,168 Canadian sequences had incomplete dates that were inferred simultaneously with internal node dates and the relaxed molecular clock, of which the greatest contributors were Ontario (n=1,338) and British Columbia (n=814), followed by Alberta (n=12) and Quebec (n=4). Time-scaled trees were resolved into binary trees by randomly resolving polytomies using the multi2di function in R package ape^42^, and branch lengths of zero were assigned a negligibly small value of the minimum non-zero branch length multiplied by 1e-8. Due to LSD2 computational limits encountered while simultaneously estimating the relaxed molecular clock while inferring incomplete tip dates, even when specifying up to 64 threads on a computing cluster, 1,348 tip dates inferred in an earlier build using data up to 15 January 2021 were input as fixed dates in LSD2 in the present analysis.

### Maximum likelihood discrete ancestral state reconstruction

Each tip state was assigned as the patient’s sampling location (except for travelers to Iran and Italy, for reasons described above), which was either Canadian province or country of sampling. We applied maximum likelihood discrete ancestral state reconstruction on the time-scaled tree using the ace function of R package ape^42^, and the highest likelihood state was pulled for each internal node. Canadian sublineages were designated as international introductions resulting in onward transmission with a minimum of two downstream sequenced cases, where a Canadian internal node was preceded by a non-Canadian internal node. It should be noted that a Canadian sublineage may comprise multiple transmission clusters or outbreaks, and could even include multiple introductions of identical viruses. We estimated the time of most recent common ancestor (tMRCA) for introduction nodes using the bootstrap time-scaled trees inferred in LSD2 via IQ-TREE 2.1.2 with a relaxed molecular clock and 100 bootstrap trees to estimate 95% confidence intervals. The tMRCA represents the first inferred transmission event resulting in a sampled Canadian sublineage. We interrogated whether any of the sublineages’ tMRCAs supported that SARS-CoV-2 was circulating prior to detection in Canada in early 2020.

### Sublineage detection lag and number of descendants analyses

Each sublineage’s detection lag was estimated as the number of days between the tMRCA and the first Canadian sample collection date (Fig. S11). The detection lag and number of descendants, respectively, between provinces were compared using a non-parametric Kruskal-Wallis rank sum test, followed by a pairwise Dunn’s rank sum test with Bonferroni p-value adjustment^46^. An alpha value of 0.05 was deemed as significant. Subsequently, we evaluated whether sublineages’ detection lag was associated with the tMRCA using multiple linear regression, as well as whether the number of descendants in each sublineage was associated with the tMRCA within a negative binomial model using the R package MASS^45^. For each model, the inclusion of province as a confounder was evaluated using anova likelihood ratio tests and Bayesian Information Criterion (BIC). Finally, the relationship between detection lag and the number of descendants was described using a negative binomial model using the R package MASS^45^ (Fig. S12).

### Origins of sampled transmission events

In addition to evaluating international importations into Canada resulting in domestic transmission (i.e., Canadian sublineages), we further evaluated the ancestral state of all Canadian tips in the phylogeny, representing all transmission events in which a sampled Canadian was the recipient. Canadian tips represent either descendants of a Canadian sublineage or singletons, which are international importation events with no further sampled downstream transmission. Notably, Geoghegan *et al*. defined a singleton as an introduction leading to one other secondary case, in contrast to an introduction with no sampled ongoing transmission^4^. However, du Plessis *et al*. defined singletons as genomes that were not assigned to any importations resulting in 2 or more sequences sampled^5^. We follow this definition, whereby singletons are sampled Canadian genomes with international transmission sources, no sampled descendants, outside of Canadian sublineages. Together, the singletons and the Canadian sublineages represent all the distinguishable, unique, and sampled importations of SARS-CoV-2 from abroad. The proportion of international importations resulting in no further transmission, i.e., a singleton, over time was stratified by season (Winter 2020: 22 December 2019 – 19 March 2020; Spring 2020: 20 March 2020 – 19 June 2020; Summer 2020: 20 June 2020 – 21 September 2020; Fall 2020: 22 September – 20 December 2020l; and Winter 2021: 21 December 2020 – 19 March 2021) and either province of introduction or origin location in order to evaluate whether there were significant temporal changes. We applied non-parametric Kruskal-Wallis tests to evaluate the hypothesis that the proportion of introductions resulting in a sublineage (versus a singleton) should decrease over time as Canada improved its testing, contact tracing, and quarantine compliance. We mapped the total and proportional contribution of all international sources to sampled transmission events by province and season (Fig. 5). Between-province sampled transmission events were summarized as a matrix (Fig. S13).

## Results

### Subsampled global sequences better reflect countries’ relative case contributions

We reduced the overrepresentation of sequences from certain countries with the highest sequencing effort (notably the UK, Australia, and Denmark) by subsampling the global sequences proportionally to each country’s contribution to global new diagnoses by month (Fig. 1a-c). Since this is a Canadian focused analysis, the contribution of subsampled Canadian sequences outweighs Canada’s overall contribution to global new diagnoses, but otherwise is representative of global pandemic dynamics (Fig. 1c). The fit of the overall relationship between countries’ monthly new diagnoses and sequences was improved following subsampling, increasing the R^2^ from 0.52 to 0.72, and yielding a final average sequence representation of 0.016 (total number sequences/total number of confirmed diagnoses) (Table S1; Fig. S3).

Since all clean Canadian GISAID sequences (n= 9,657) were kept in the subsampling process to maximize our ability to detect international importations, there remain discrepancies between Canadian provinces’ contributions of new diagnoses (among provinces with available sequences) and contributions of sequences over time (Fig. 1d-e). However, the overall sequence representation relative to new diagnoses was comparable across provinces (Fig. 1f), with a median sequence representation of 0.014 (minimum= 0.010, Quebec; maximum= 0.29, Nova Scotia). Gaps in the availability of sequences over time varied greatly by province (Table S2) and is a caveat to our analysis. There are no sequences representing British Columbia, New Brunswick, Newfoundland and Labrador, or Nova Scotia for summer 2020 (and only 1 sequence from Alberta). Quebec has not deposited any sequences corresponding to collection dates after September 2020, and there are zero sequences sampled in fall 2020 from Alberta, New Brunswick, Newfoundland and Labrador, and Nova Scotia. Additionally, there were no sequences available from any cases in Yukon, Northwest Territories, Nunavut, Saskatchewan, or Prince Edward Island, despite representing 3.4% COVID-19 cases in Canada (Table S3).

### Diversity of SARS-CoV-2 in Canada

We estimated that the root age of the subsampled SARS-CoV-2 global phylogeny with a Canadian focus was 09 December 2019 (95% CI: 27 October – 10 December**) (Fig. 2)** and exhibited a lognormal relaxed clock rate of 2.66e-4 (2.61e-4, 2.70e-4) substitutions/site/year after having specified a prior of 0.2 standard deviation. This contrasts notably with the strict clock rate that we inferred in Tempest, which after removing temporal outliers was estimated as 7.47e-4 substitutions/site/year (Fig. S4d). Notably, these were both estimated on trees inferred from masked alignments.

**Fig. 2.**
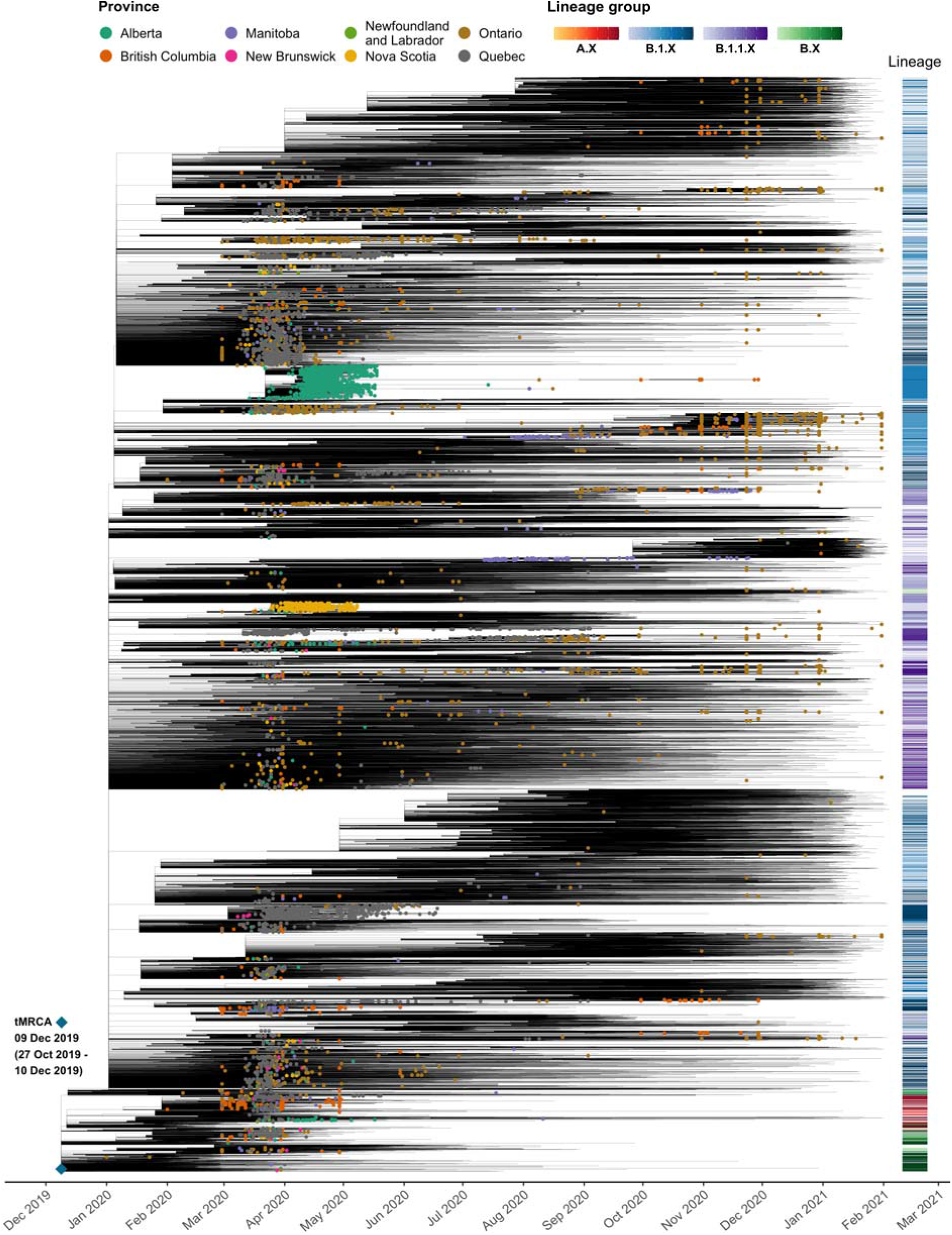
Canada in the context of a global SARS-CoV-2 time-scaled maximum likelihood phylogeny. A representative approximate maximum likelihood tree was generated using FastTree2, rooted, cleaned of temporal outliers (final n=49,236), then converted to a time-scaled tree using LSD2 with a relaxed clock 100 bootstraps for confidence intervals on date estimations. Canadian sequences are colored by province and enlarged. Pango lineages are shown on the right y-axis.

The high diversity of SARS-CoV-2 lineages sequenced in Canada illustrates the role of international transmission in the context of the global pandemic. A total of 79 unique Pango lineages were detected in the clean Canadian sequences publicly available at the time of analysis, however, since the same lineage may have been introduced multiple times independently, examining the number of lineages over time in Canada does not directly address whether these viruses were the result of singular or multiple introduction events.

### Diverse origins of Canadian sublineages

By applying discrete ancestral state reconstruction upon time-scaled phylogenies, we conservatively estimated that there were at least 402 SARS-CoV-2 introductions resulting in ongoing transmission into Canada by February 2021 (Fig. 3). Additionally, we identified 1,380 singleton importations that did not result in further sampled transmission and were not within a Canadian sublineage. Since the 9,657 clean samples represent 1.2% of the 817,163 confirmed diagnoses in Canada on 11 February, there could have been upwards of 33,500 sublineages identified had all confirmed diagnoses been sequenced, assuming that samples were sequenced randomly. If we further account for unascertained cases and assume a conservative mean case detection rate of 0.42^47^, this upper estimate doubles. While these assumptions are likely imprecise, our estimates are nonetheless conservative.

**Fig. 3.**
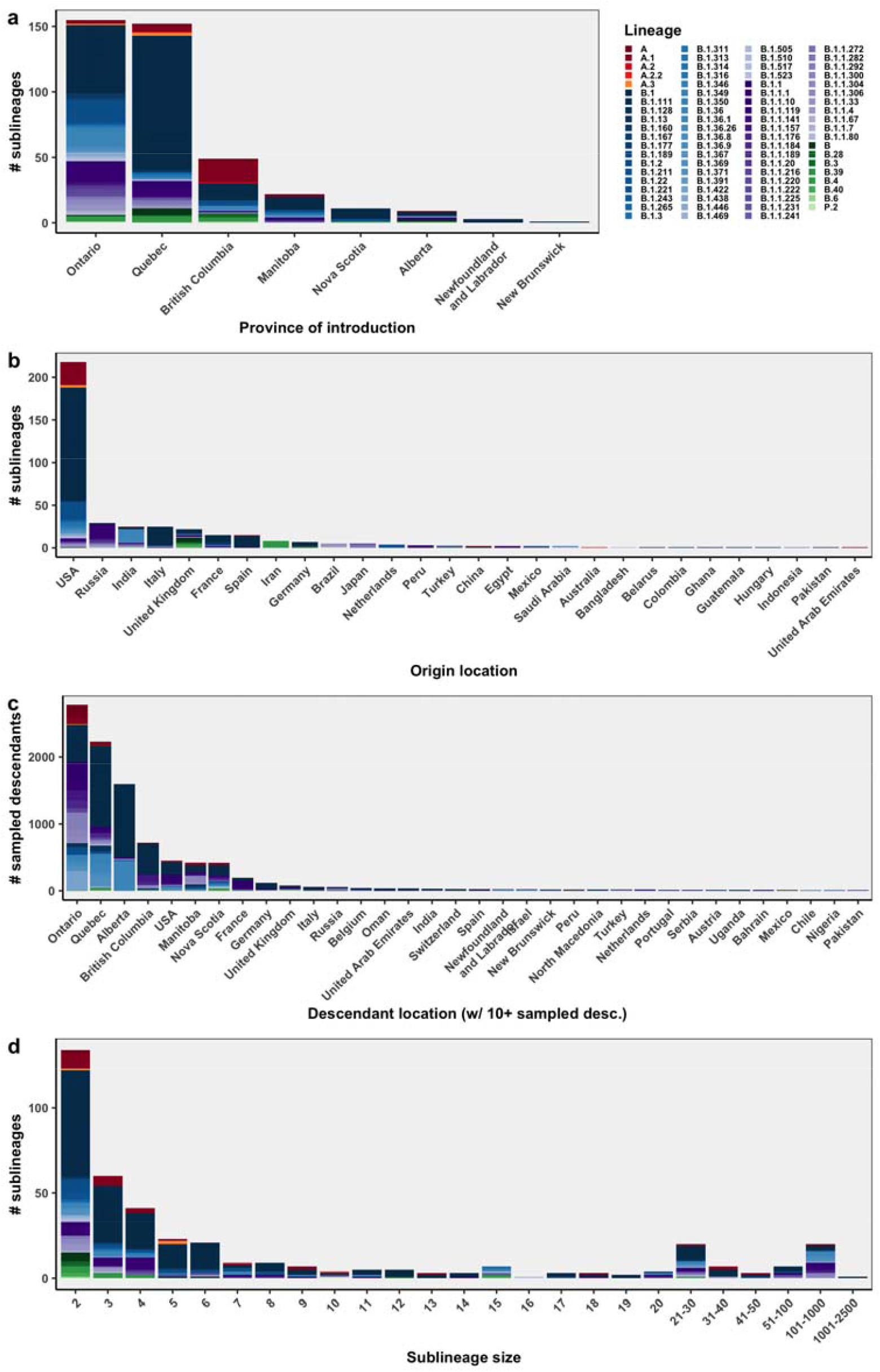
The overall representation of Pango lineages among Canadian sublineages, defined as inferred introductions of SARS-CoV-2 into Canada from abroad that resulted in downstream sampled transmission. Canadian sublineages are summarized by their a) province of introduction b) origin location, c) descendant locations (for locations with more than ten sampled descendants) and d) size, i.e. number of sampled descendants.

The large majority of sublineages were supported by high likelihoods (>0.9) in both introduction and parent nodes (79%) (Fig. S5). The majority of introductions were into Ontario (n=155), followed by Quebec (n=152), British Columbia (n=49), Manitoba (n=22), Alberta (n=9), Nova Scotia (n=11), Newfoundland and Labrador (n=3), and New Brunswick (n=1) (Fig. 3a). B.1 was the most frequently introduced lineage (n=191); however we note it includes a broad diversity of sequences. Additionally, there were more than ten introductions of A.1 (n=25), B.1.1.119 (n=23), B.1.2 (n=21), and B.1.36 (n=14) (Fig. S6). There was only one introduction of a VOC (B.1.1.7) resulting in two sampled descendants; however, since we completed our analysis, this number has increased substantially following the completion of this analysis.

The proportion of confirmed diagnoses that were sequenced was comparable across provinces (Fig. 1f); however, seasonally, there were large gaps in sequence representation from particular provinces (i.e. no sequences available from British Columbia in summer 2020) (Fig. 1e; Table S2) that lend some uncertainty to these results. While cases in Canada were low in summer 2020 (Table S3), they were not zero. Low sequence representation likely led to underestimations of the number of introductions, but could also have prevented us from connecting sequences between provinces that were linked in the transmission chain.

On the origin of Canadian sublineages (Fig. 3b), only two introductions were inferred to have originated directly from China, whilst 54% (n=218) of sublineages originated from the USA, followed by Russia (n=29), Italy (n=25), India (n=25), UK (n=22), Spain (n=15), France (n=15), and Iran (n=8) among others. We identified 9,742 downstream sampled descendants of Canadian sublineages isolated from 77 other countries, representing all continents except Antarctica (Fig. 3c). While the majority of descendants were sampled within Canadian provinces (n=8,210), this should be interpreted with caution as the global sequences were heavily subsampled – therefore sampled descendants are vastly underestimated. Sublineage sizes (number of sampled descendants) varied widely (Fig. 3d) and the largest sublineage had 2,137 sampled descendants and was of the B.1 lineage.

### Sublineage introduction rates over time

Using the time-scaled phylogenies, we estimated the time of the most recent common ancestors (tMRCA) corresponding to introduction nodes. The tMRCA is an approximation of the date of the first transmission event resulting in a downstream sampled descendant following introduction of the virus into Canada, which can differ from the index case by one or more generations. By tracking the tMRCA of sublineages over time, we can decipher the temporal dynamics of SARS-CoV-2 importations into Canada (Fig. 4). Equivalent plots with sublineages’ first Canadian sample collection dates are in Fig. S9. The majority of sublineages were introduced in February and March of 2020 across all provinces, but most notably into Quebec, which had a maximal weekly rate of 32.7 new sublineages per week on 21 March 2020, without including singletons. Correspondingly, a maximum of 31.1 new sublineages were introduced from the USA on 22 March. Ontario reached its peak a week later on 29 March, wherein the rolling weekly introduction rate peaked at 11 new sublineages per week. It is worth noting that over this period, a large number of Canadians were repatriated from around the world and this likely contributed to a delay in reduction of the importation rates. Subsequently, new sublineage introduction rates across most provinces dropped sharply; for instance, comparing the mean weekly introduction rates from 1 week and 4 weeks after travel restrictions were enacted, Quebec British Columbia, and Ontario respectively had 18.5, 2.5, and 1.9 times fewer new sublineage introductions. Despite these reductions, introductions were not eliminated. Throughout the summer and fall of 2020, we detected ongoing introductions into Quebec, Ontario, Manitoba, and British Columbia imported from the USA, India, Russia, and other nations. Again, we must emphasize that our ability to detect importation is directly proportional to the availability of sequences. For instance, there were no available sequences from British Columbia in the summer of 2020, and the only sequences we have from New Brunswick and Alberta are from the spring of 2020. The importation rate increased in Ontario (the province with the most consistently deposited sequences and the most incomplete sample dates) in September 2020 and again at the end of November 2020 – on 27 November, there were on average 8.1 new sublineages introduced into Ontario per week. Tracking the first Canadian sample date (Fig. S9) instead of the date of the most recent common ancestor (Fig. 4), revealed a similar pattern with a delayed peak in sampled introductions due to the detection lag. The most recent sublineage introduction we detected was the B.1.1.7 lineage into Ontario from UK in the last week of December, resulting in two sampled descendants.

**Fig. 4.**
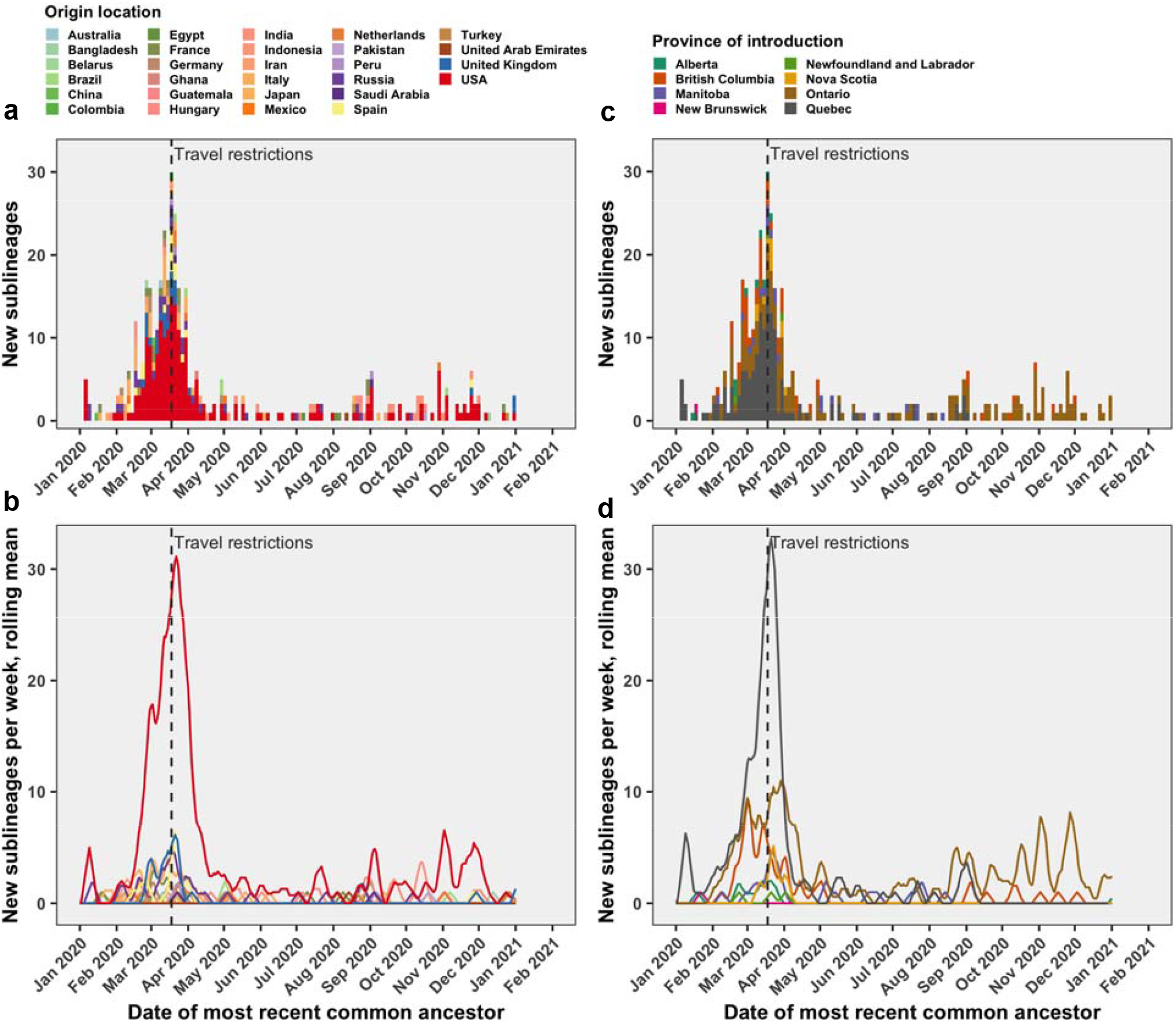
Following the implementation of restrictions on non-essential travel on 18 March 2020 (dashed grey line), introductions resulting in ongoing transmission decreased, but were not eliminated. Equivalent plots using the First Canadian sample collection date (Fig. S9).

### Sublineage detection lags and number of descendants analyses

We estimated the detection lag as the number of days between the tMRCA and the first Canadian sample collection date among sublineage descendants (Fig. S11). The importation date of virus sublineages first arriving in Canada is more challenging to infer and precedes the tMRCA by an importation lag4. By applying a Kruskal-Wallis test, we identified that there were significant differences between sublineages’ detection lags between provinces (p-value = 0.016) (Fig. S12a). A further pairwise Dunn’s test revealed that the only significant difference was between Ontario and British Columbia (p=0.019); the median detection lag in Ontario was 13 while it was only 3 in British Columbia. Upon investigating whether detection lag was associated with sublineages’ tMRCA, adjusting for province of introduction significantly improved the goodness-of-fit as assessed by a likelihood ratio test although there was no support for effect modification. Thus, we found a modest, but significant effect that for every 30 days later in the year the tMRCA occurred, the detection lag was on average 2.0 days smaller (0.93 - 0.31), adjusted for province (Fig. 12b; Table S4). Subsequently, we evaluated whether sublineages’ size was associated with the tMRCA. In exploratory analyses, we found no significant differences of sublineage sizes between provinces (Kruskal-Wallis: p-value = 0.08). However, we found a significant relationship between the tMRCA and number of sampled descendants in a negative binomial model without adjusting for province (Fig. 12c; Table S5). Upon identifying decreasing trends of both sublineage size and detection lag over time, we evaluated their relationship directly in a negative binomial model, adjusting for province. We found that that longer detection lags were significantly associated with larger sublineages (p<0.001), such that for every 10 days additional lag, there were on average 10.2 additional sampled descendants, adjusted for province (Fig. 12d; Table S6).

### Relative transmission source contributions and singletons

To further evaluate the hypothesis that travel restrictions reduced, but did not eliminate, international importations, we queried the parental node state for all Canadian tips to discern the likely origins of sampled tips, to identify singletons, and to quantify relative contributions of sampled transmission attributable to either: within-province, interprovincial, USA, or other international. In contrast to our sublineage analyses above, this analysis focused solely on transmission events immediately preceding a sampled tip.

There were 1,380 singleton genomes, which were likely importations from international sources without sampled descendants that were not in a Canadian sublineage. Upon removing the criteria to be outside a sublineage, an additional 7 singletons appeared. It is possible that singletons were designated as such only because their descendants were not included in the subsample (or ever sampled). The distributions of tips’ locations (transmission recipient) and inferred locations of origin (transmission source) over time (Fig. S14; S15) largely mimic the trends seen among introductions resulting in domestic transmission (Fig. 4), with a peak in importations in late March, but ongoing contributions of low-level international and interprovincial transmission throughout 2020.

We aggregated the results by province and by season in order to evaluate the relative contributions of sampled transmission attributable to within-province, interprovincial, USA, and other international. As the border closure occurred on 18 March 2020, a few days before the advent of spring 2020, comparing winter and spring 2020 permits the analysis of whether that intervention was effective in reducing importations. The contribution of within-province transmission steadily increased over time for all provinces, coinciding with a reduction, but not an elimination, in the relative contributions of USA, other international, and interprovincial transmission sources over time (Fig. S16). However, if we look at the total number of sampled transmission events traced to international origins instead of the proportion (Fig. 5), there was an increase in the number of detected transmission events traced to USA or other international sources between winter and spring 2020 for all provinces. It was not until the summer of 2020 that there was a marked reduction in international transmission (in provinces with samples available, excluding British Columbia, Alberta, and the Maritime provinces). Again, this finding is limited by the availability of sequences across provinces. Ontario and Manitoba were the only provinces to consistently deposit sequences across all seasons and therefore have the highest confidence. Results from fall 2020 and winter 2021 should be interpreted cautiously as the lag between sample collection and sequence deposition on GISAID has been variably lengthy^48^

**Fig. 5.**
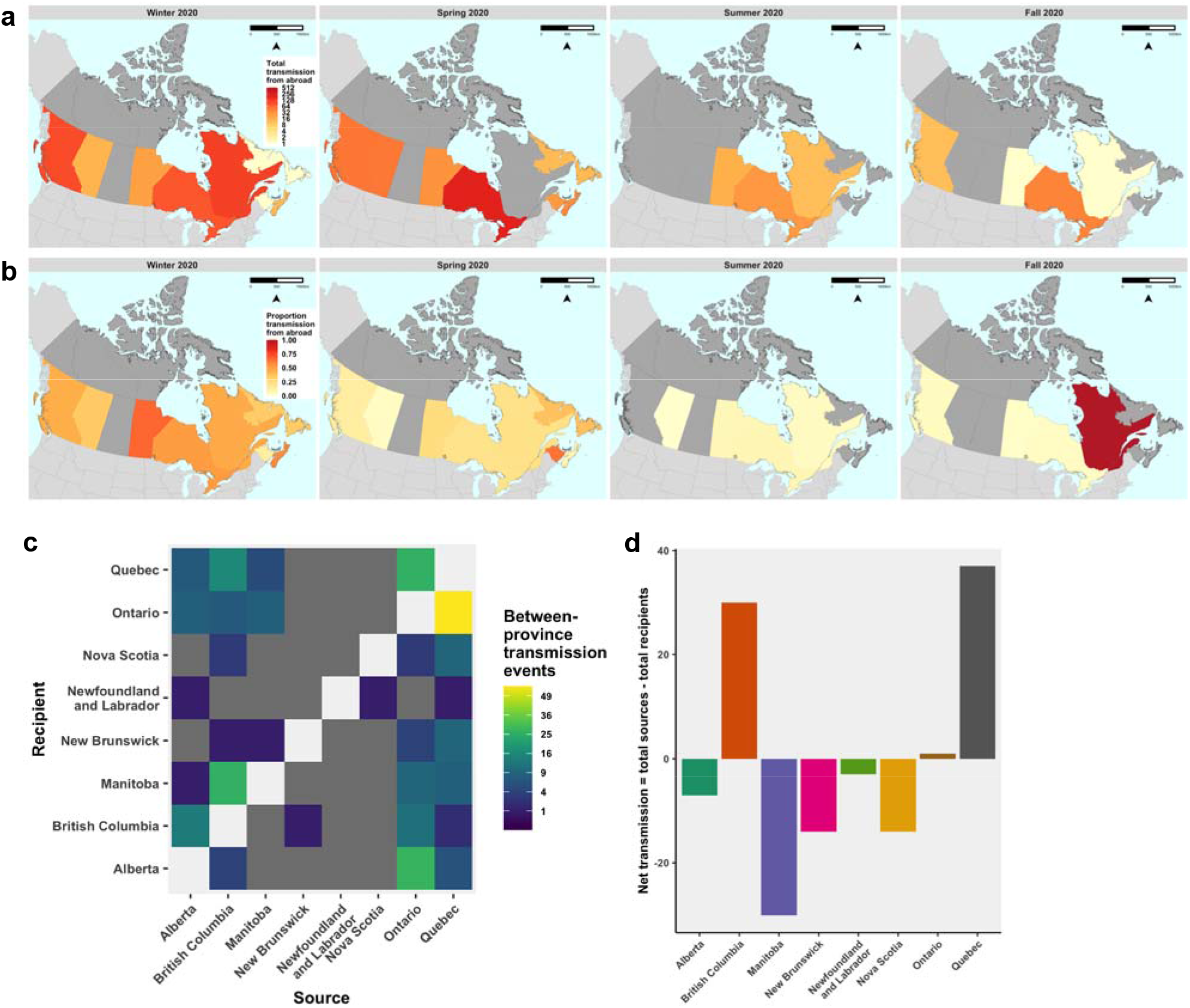
Contribution of international transmission sources among all sampled tips across Canadian provinces and by season. c) Interprovincial transmission was quantified among Canadian tips with Canadian origin from outside province. Diagonal cells for intraprovincial transmission were omitted; dark grey cells indicate zero values. d) The net difference between a province being either the source or recipient across sampled transmissions was compared.

Interprovincial transmission was further evaluated by comparing inferred transmission origins among Canadian sequences with a Canadian parental node from another province (Fig. 5), where it is evident that Ontario and Quebec were the greatest sampled sources of interprovincial transmission, and British Columbia acted as a notable transmission source to Manitoba in spring 2020 (Fig. 5c). For each province, the net transmission was calculated as the difference in sampled transmission events where each province was inferred as the source or the recipient. Quebec and British Columbia were the greatest net sources of transmission (Fig. 5d), however this result is uncertain due to poor data availability. Specific province comparisons revealed contrasting source-sink dynamics; for example, there were more inferred transmission events from Alberta to British Columbia than from British Columbia to Alberta. The extent of mixing between Canadian provinces is illustrated by numerous sublineages comprised of sequences from multiple provinces (Fig. 2). Finally, we evaluated whether the proportion of all importations resulting in domestic transmission (i.e. a Canadian sublineage) rather than a singleton changed significantly over time. By binning these events by season and summing by province of introduction (Fig. S13a) or origin location (Fig. S13b), we were able to compare the distributions of proportions using a non-parametric Kruskal-Wallis test (p=0.58). The winter 2021 season was removed from the analysis due to low sequence availability. Contrary to our expectations, the proportion of importations resulting in a sublineage did not significantly change over the course of 2020.

## Discussion

Our analyses revealed that Canadian travel restrictions imposed in March 2020 to limit the COVID-19 burden reduced but did not eliminate international seeding of SARS-CoV-2 into Canada throughout 2020. Furthermore, unhampered interprovincial travel of infectious Canadians further contributed to ongoing transmission of SARS-CoV-2 between provinces. There were significant reductions in sublineages’ detection lag, adjusted for province, and number of sampled descendants over time, suggesting that contact tracing and outbreak control did somewhat improve over the course of the pandemic. However, the proportion of importations resulting in ongoing domestic sublineages did not change over time, indicating that more could have been done to stop new viral introductions from igniting outbreaks. Although non-essential international travel was restricted on 18 March 2020, repatriation of Canadians and domestic travel contributed to the continued spread of SARS-CoV-2 in Canada.

Discrete ancestral trait reconstruction is sensitive to sampling bias, causing ancestral nodes to be more likely to share the states predominant among tips. We reduced sampling bias by subsampling sequences by countries’ proportional contribution to global diagnoses in each month. This method, however, is sensitive to differences in countries’ contact-tracing efforts, testing strategies, and case reporting. Further, while subsampling can effectively reduce overrepresentation of geographies that contributed a higher proportion of sequences than proportion of new diagnoses, such as the UK, this method does not directly improve representation of geographies that are undersampled in the sequence database. Since we had a priori knowledge from Worobey *et al*.^1^ that Iran and Italy were undersampled early in the COVID pandemic, yet played a key role as sources of infection, we were able to designate sequences from individuals who had travelled to those countries as originating from those countries, this was relatively ad hoc and not possible in the absence of travel data. Incorporating individuals’ travel history when available, as demonstrated by Lemey *et al*.^49^, or flight volume data^5^ could help to improve representation of otherwise undersampled geographies. Our analyses are limited to that which was measured and shared publicly.

Differences in sequence sharing across Canadian provinces over time added uncertainty to our results. Large gaps in publicly shared genomes, where for instance no sequences from British Columbia are available for summer 2020 (Table S1), make these analyses challenging to execute and interpret appropriately, particularly when it comes to comparing provinces’ relative international importations as well as temporal trends for a given province. We expect that our ancestral reconstruction was more robust to origin location than provinces of introduction due to differences in provinces’ sequence deposition. Additionally, the deposition of Canadian SARS-CoV-2 sequences with incomplete sampling dates presented a computational challenge that could have been abrogated by open data sharing. While protecting patient privacy and confidentiality is important, it is unlikely that release of a sampling date associated with a viral genetic sequence would result in re-identification of any individual. The added computational time and parallelization required to simultaneously infer incomplete sample dates for thousands of tips while estimating a relaxed molecular clock during time-scaled tree conversion using LDS2 was not insignificant, and added unnecessary uncertainty into the phylogeny and clock estimation. Meanwhile, nearly all the other nations in the world sharing SARS-CoV-2 sequences have shared full sample collection dates, and Japan recently added >15,000 complete dates to previously submitted sequences with incomplete dates on GISAID. Canada was ranked 83^rd^ compared to other nations for the proportion of genomes with complete sample collection dates (around 1/3) and 80^th^ in the average delay between sample collection and release of genome data, with an average of 168 days passed between sample collection and sequence release^48^. Overall, these inconsistencies create unnecessary burdens to interpreting patterns across provinces and time. Improvements in reporting sequence dates and closing the gap between sequencing and reporting would enhance public health in Canada during this, but also future, pandemics.

Seeing as the Canadian virus genomes available only represent a small proportion of diagnosed cases (1.05% overall) and an even smaller proportion when taking into consideration undiagnosed cases^47^, our estimates of the number of importations are severely underestimated. Larger sublineages are more likely to be well-represented than smaller sublineages, and a proportion of the singletons were likely part of local transmission networks that were unsampled. Furthermore, separate chains of transmission with identical viral genomes could lead to grouping sequences that were in fact independent introductions that could not be disentangled, further underestimating the number of introductions. An additional consideration in the interpretation of sublineages as independent introductions is the possibility that seemingly distinct sublineages were the result of a single introduction that circulated within Canada for enough generations to have accumulated at least one mutation to distinguish them before they were detected by sequencing efforts. This would lead to an overestimate of the number of independent introductions.

Another source of sampling bias worth considering is that sequences were more likely to be generated for patients with low cycle threshold values, due to a higher viral load. This could bias the analysis towards viruses from individuals who were sampled at the peak of their infectious period or who experienced more severe symptoms, whereas individuals sampled too early or late in their infection or who had a low viral load throughout might not be represented. If we assume that individuals with a higher viral load are more infectious^50,51^, then their contribution to the transmission network would be greater, justifying their relative overrepresentation in genomic epidemiological analyses.

Overall, it was not surprising that the USA was the greatest source of SARS-CoV-2 importations into Canada considering our shared land border – the longest in the world - and close trade relationships. What was concerning, however, was our inference of ongoing importations from the USA even after restricting non-essential international travel. In light of wide variations in provinces’ COVID intervention stringency and duration^21^, it is unsurprising that many Canadians and non-residents continued to travel internationally throughout 2020 and early 2021^52^. Additional ongoing importations from the UK, Russia, and India, among others, further indicates that there is room for improvement to better enforce quarantine, testing, and contact tracing upon return to Canada for essential workers and non-essential travellers alike. One consideration would be to enforce (affordable and subsidized) mandatory 14-day hotel quarantine upon first landing, as in Australia and New Zealand, removing the possibility of transmission during domestic connecting flights, public transport, and interactions with family and community members while in self-isolation. While the majority of importations were inferred to have been introduced into Ontario, this is partially attributable to the consistent sequence deposition over time from Ontario. It is also worth noting that the majority of Canadian sequences deposited into the public domain without incomplete sample collection dates came from Ontario. By 11 February 2021, Quebec had deposited a single sequence since September 2020 and Alberta had deposited one since May 2020. Based on provincial population sizes alone, we would expect most importations into Ontario and Quebec (with populations of 14.8M and 8.6M, respectively) than into British Columbia (5.2M), Alberta (4.5M), or other provinces and territories with less than 1.5M^53^.

Consistent with the findings from the UK and NZ, which found that transmission lineages generally became smaller^5^ over time, we found a significant reduction in the number of sampled descendants of Canadian sublineages over time. We note, however, that this result could be affected by the delay in depositing sequence data for more recent sublineages. We additionally found that the detection lag was significantly lower over time, adjusting for province of introduction, and that the longer the detection lag, the more sampled descendants there were. This is intuitive as interruption of further transmission through contact tracing, case isolation, and testing, is dependent upon detecting the transmission chain in the first place. Our results therefore corroborate the importance of timely public health interventions.

Our analysis of whether there were temporal changes in the proportion of importations resulting in ongoing transmission, as opposed to singletons, was somewhat underpowered to detect any significant changes over time. Within Ontario, the proportion increases into fall 2020, serving as further evidence that enforcement and/or compliance of the 14-day quarantine upon returning to Canada did not improve over time. Notably, singletons may simply reflect a lack of sequences representing downstream domestic transmission, not true evolutionary dead ends. Therefore, we expect that the proportion of importations resulting in ongoing transmission are underestimates, particularly in seasons where few to no sequences are available from certain provinces.

We anticipate that the recent rise of VOCs in Canada has been the result of multiple introductions of B.1.1.7, P.1, B.1.351, and we look forward to these sequences being shared with complete dates, empowering Canadian researchers to contribute to our understanding and response to the ongoing COVID-19 pandemic.

## Conclusions

Our phylogeographic analyses illuminated the extent to which the COVID-19 pandemic in Canada was perpetuated by ongoing international importations and interprovincial transmission throughout 2020. Although travel restrictions enacted in March 2020 reduced the importation rate and proportion of transmission from abroad across all Canadian provinces, SARS-CoV-2 introductions from the USA, India, Russia, and other nations were detectable through the summer and fall of 2020. We provide evidence supporting that public health responses led to a modest reduction of imported sublineages’ size and detection lag over time.

Understanding SARS-CoV-2 transmission dynamics in response to public health interventions can inform their effectiveness in diminishing ongoing transmission. Sharing viral genome sequences linked with the time and place of sampling in a timely manner is of utmost importance for public health; doing so enables epidemic surveillance for new and already described variants of concern, analyses to support contact tracing, and the inference of the timing and routes of SARS-CoV-2 migration, which can inform the effectiveness of public health interventions.

## Data Availability

All the viral genetic sequences and associated metadata used in this analysis are publicly available on GISAID, where open access to data is provided free-of-charge to all individuals that agreed to identify themselves and agreed to uphold the GISAID sharing mechanism governed through its Database Access Agreement. A list of included sequence identifiers is included in Appendix 1. The code can be shared upon request.

## Acknowledgements

We are especially thankful to the efforts of the CanCOGeN (Canadian COVID-19 Genomics Network) VirusSeq and the CPHLN (Canadian Public Health Laboratory Network) for making data from Canada publicly available on GISAID. We are also very grateful to all the contributing laboratories within Canada and internationally that generously deposited sequences and metadata on GISAID (Supplementary Appendix 1).

## Supplementary Material

**Appendix 1**. An acknowledgment of contributing laboratories who generated viral genetic sequences and uploaded them to GISAID with metadata.

**Table S1.** The number of available clean global and Canadian sequences in comparison to the number of sampled global and Canadian sequences by month.

| Month | Clean Global | Sampled Global | Clean Canadian | Sampled Canadian |
| --- | --- | --- | --- | --- |
| Dec 2019 | 22 | 22 | 0 | 0 |
| Jan 2020 | 491 | 491 | 4 | 4 |
| Feb 2020 | 1,214 | 1,214 | 7 | 7 |
| Mar 2020 | 42,194 | 3,500 | 2,533 | 2,533 |
| Apr 2020 | 41,789 | 3,500 | 3,065 | 3,065 |
| May 2020 | 20,575 | 3,500 | 912 | 912 |
| Jun 2020 | 22,921 | 3,500 | 275 | 275 |
| Jul 2020 | 24,095 | 3,500 | 257 | 257 |
| Aug 2020 | 27,332 | 3,500 | 374 | 374 |
| Sep 2020 | 29,661 | 3,500 | 251 | 251 |
| Oct 2020 | 49,882 | 3,499 | 122 | 122 |
| Nov 2020 | 57,971 | 3,499 | 1,277 | 1,277 |
| Dec 2020 | 69,489 | 3,499 | 475 | 475 |
| Jan 2021 | 68,612 | 3,499 | 105 | 105 |
| Feb 2021 | 120 | 120 | 0 | 0 |
| **TOTAL** | **456,368** | **40,343** | **9,657** | **9,657** |

**Table S2.** The number of included SARS-CoV-2 sequences by Canadian province and month.

| Month | Alberta | British Columbia | Manitoba | New Brunswick | Newfoundland and Labrador | Nova Scotia | Ontario | Quebec |
| --- | --- | --- | --- | --- | --- | --- | --- | --- |
| Jan 2020 | 0 | 0 | 0 | 0 | 0 | 0 | 4 | 0 |
| Feb 2020 | 0 | 2 | 0 | 0 | 0 | 0 | 5 | 0 |
| Mar 2020 | 140 | 245 | 114 | 39 | 31 | 85 | 431 | 1,448 |
| Apr 2020 | 1,227 | 295 | 30 | 12 | 4 | 347 | 449 | 701 |
| May 2020 | 307 | 34 | 4 | 0 | 0 | 38 | 130 | 399 |
| Jun 2020 | 0 | 0 | 6 | 0 | 0 | 0 | 213 | 56 |
| Jul 2020 | 1 | 0 | 81 | 0 | 0 | 0 | 61 | 114 |
| Aug 2020 | 0 | 0 | 138 | 0 | 0 | 0 | 113 | 123 |
| Sep 2020 | 0 | 0 | 25 | 0 | 0 | 0 | 178 | 48 |
| Oct 2020 | 0 | 89 | 0 | 0 | 0 | 0 | 33 | 0 |
| Nov 2020 | 0 | 203 | 94 | 0 | 0 | 0 | 980 | 0 |
| Dec 2020 | 0 | 1 | 0 | 0 | 0 | 0 | 473 | 1 |
| Jan 2021 | 0 | 0 | 0 | 0 | 0 | 0 | 105 | 0 |
| **TOTAL** | **1,675** | **869** | **492** | **51** | **35** | **470** | **3,175** | **2,890** |

**Table S3.** The number of SARS-CoV-2 confirmed diagnoses by Canadian province and month. No sequences were available from provinces highlighted in light blue, which collectively accounted for 3.4% of confirmed diagnoses.

| Month | Alberta | British Columbia | Manitoba | New Brunswick | Newfoundland and Labrador | Nova Scotia | Ontario | Quebec | Northwest Territories | Nunavut | Prince Edward Island (PEI) | Saskatchewan | Yukon |
| --- | --- | --- | --- | --- | --- | --- | --- | --- | --- | --- | --- | --- | --- |
| Jan 2020 | 0 | 1 | 0 | 0 | 0 | 0 | 0 | 0 | 0 | 0 | 0 | 0 | 0 |
| Feb 2020 | 0 | 6 | 0 | 0 | 0 | 0 | 5 | 0 | 0 | 0 | 0 | 0 | 0 |
| Mar 2020 | 754 | 963 | 103 | 70 | 152 | 147 | 1,958 | 4,162 | 1 | 0 | 21 | 184 | 5 |
| Apr 2020 | 4,601 | 1,142 | 172 | 48 | 106 | 800 | 14,221 | 23,376 | 4 | 0 | 6 | 205 | 6 |
| May 2020 | 1,655 | 461 | 20 | 14 | 3 | 109 | 11,672 | 23,521 | 0 | 0 | 0 | 257 | 0 |
| Jun 2020 | 1,098 | 343 | 30 | 33 | 0 | 6 | 7,209 | 4,399 | 0 | 0 | 0 | 139 | 0 |
| Jul 2020 | 2,735 | 725 | 90 | 5 | 5 | 7 | 4,141 | 3,854 | 0 | 0 | 9 | 534 | 3 |
| Aug 2020 | 3,059 | 2,149 | 799 | 21 | 3 | 16 | 3,100 | 3,180 | 0 | 0 | 8 | 300 | 1 |
| Sep 2020 | 4,160 | 3,348 | 779 | 9 | 5 | 3 | 9,401 | 11,796 | 0 | 0 | 15 | 294 | 0 |
| Oct 2020 | 10,183 | 5,595 | 3,730 | 143 | 17 | 21 | 24,020 | 31,728 | 5 | 0 | 5 | 1,231 | 8 |
| Nov 2020 | 29,932 | 18,505 | 11,102 | 158 | 47 | 196 | 40,762 | 36,355 | 5 | 181 | 8 | 5,420 | 24 |
| Dec 2020 | 43,451 | 18,752 | 7,875 | 98 | 52 | 181 | 65,667 | 60,270 | 9 | 85 | 24 | 6,785 | 13 |
| Jan 2021 | 22,580 | 15,670 | 4,864 | 657 | 18 | 94 | 86,052 | 59,942 | 7 | 28 | 15 | 8,515 | 10 |
| Feb 2021 | 9,296 | 11,602 | 2,295 | 174 | 580 | 61 | 32,605 | 25,157 | 11 | 63 | 21 | 4,783 | 2 |
| **TOTAL** | **133,504** | **79,262** | **31,859** | **1,430** | **988** | **1,641** | **300,813** | **287,740** | **42** | **357** | **132** | **28,647** | **72** |

**Fig. S1.**
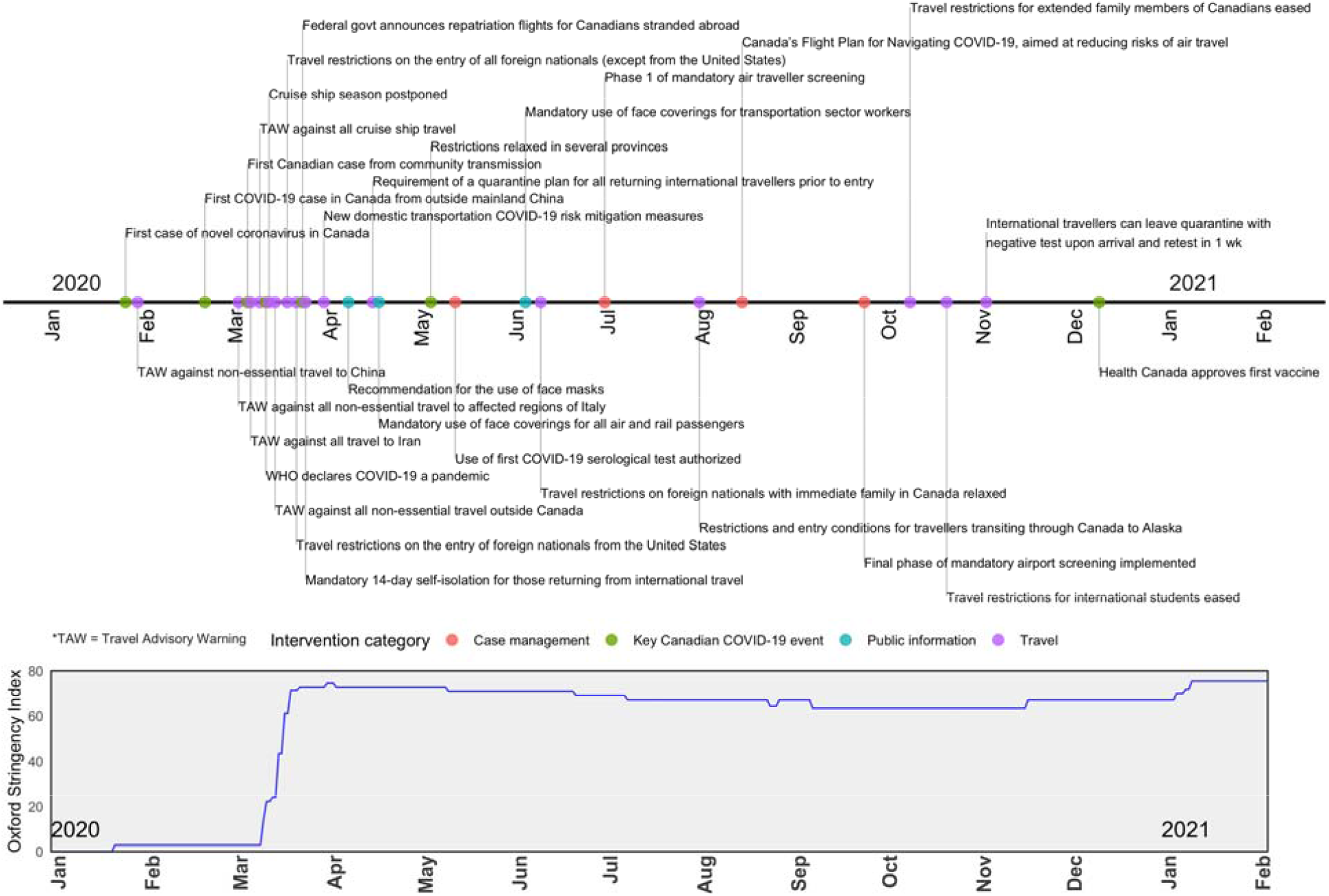
Timeline of COVID-19 in Canada in 2020, highlighting key events and interventions, as well as the Oxford Stringency Index..

**Fig. S2.**
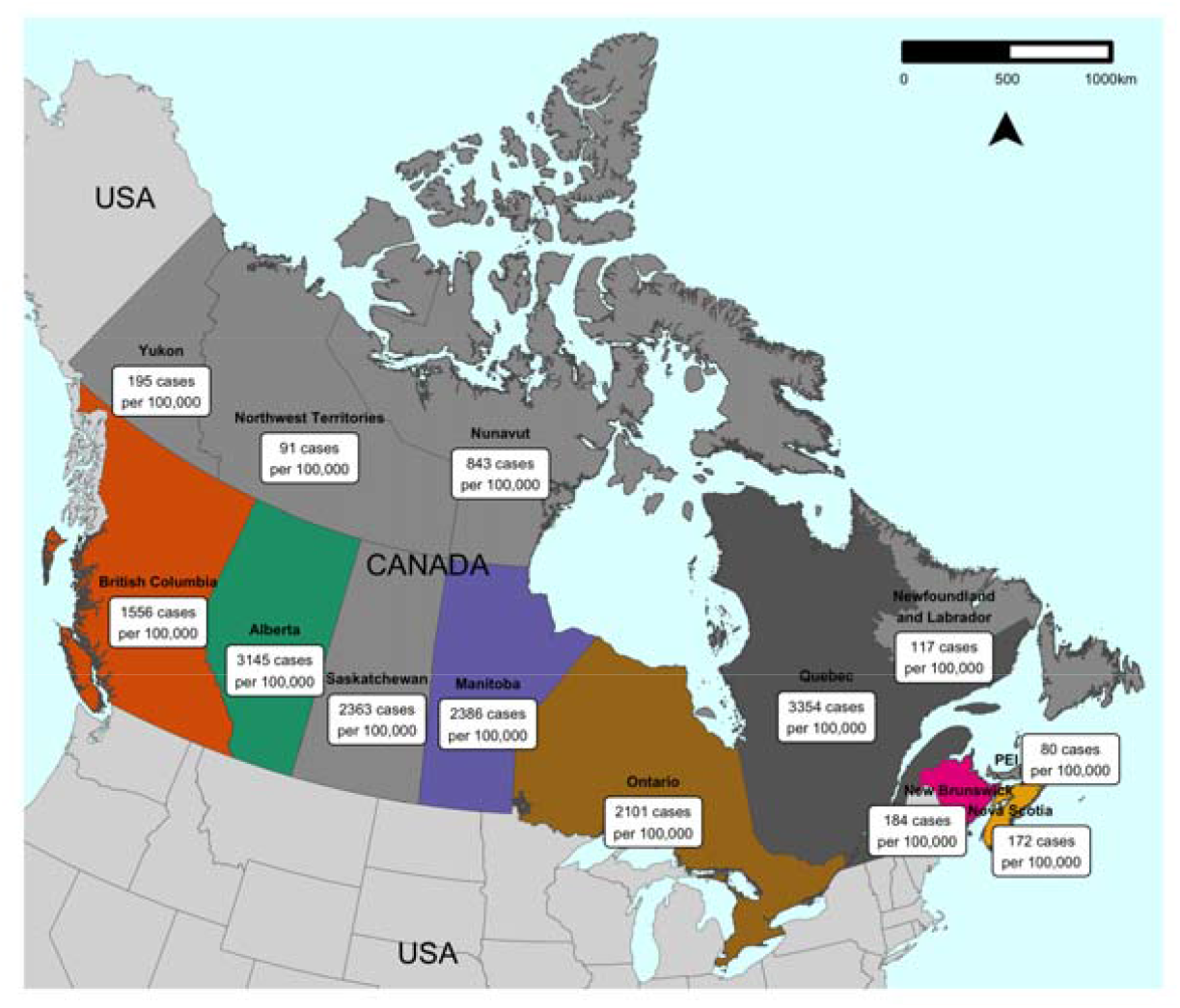
Map of Canada, labelled with provinces and territories, as well as the cumulative incidence (total new diagnoses/population in 2016 census*100,000) of COVID-19 cases on the day of data download, 11 February 2021. Colors reflect which provinces had clean viral sequences available on GISAID for the study, while others are shaded in light grey.

**Fig. S3.**
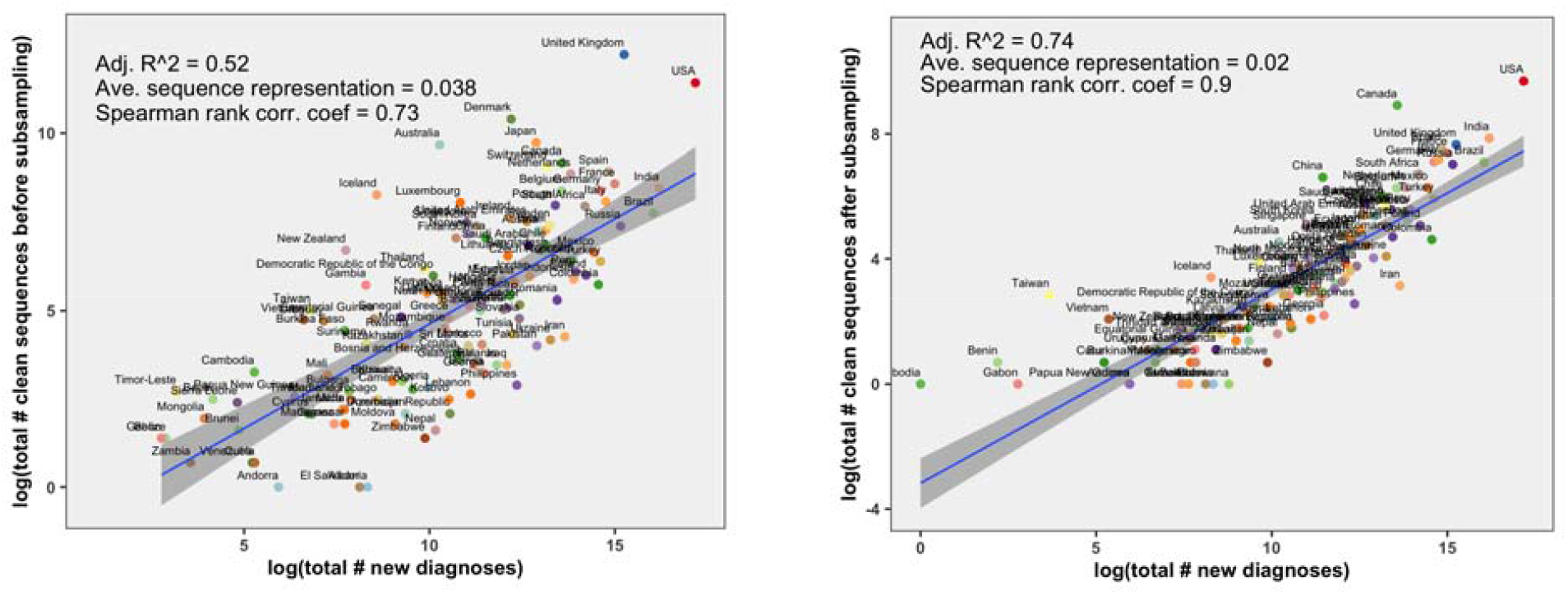
The relationship between total number of diagnoses and total number of representative sequences in the dataset by country a) before and b) after subsampling global sequences relative to their case contributions. The average sequence representation is the total number of sequences from a country divided by the total number of COVID-19 diagnoses. This analysis excludes countries for which there were zero sequences available.

**Fig. S4.**
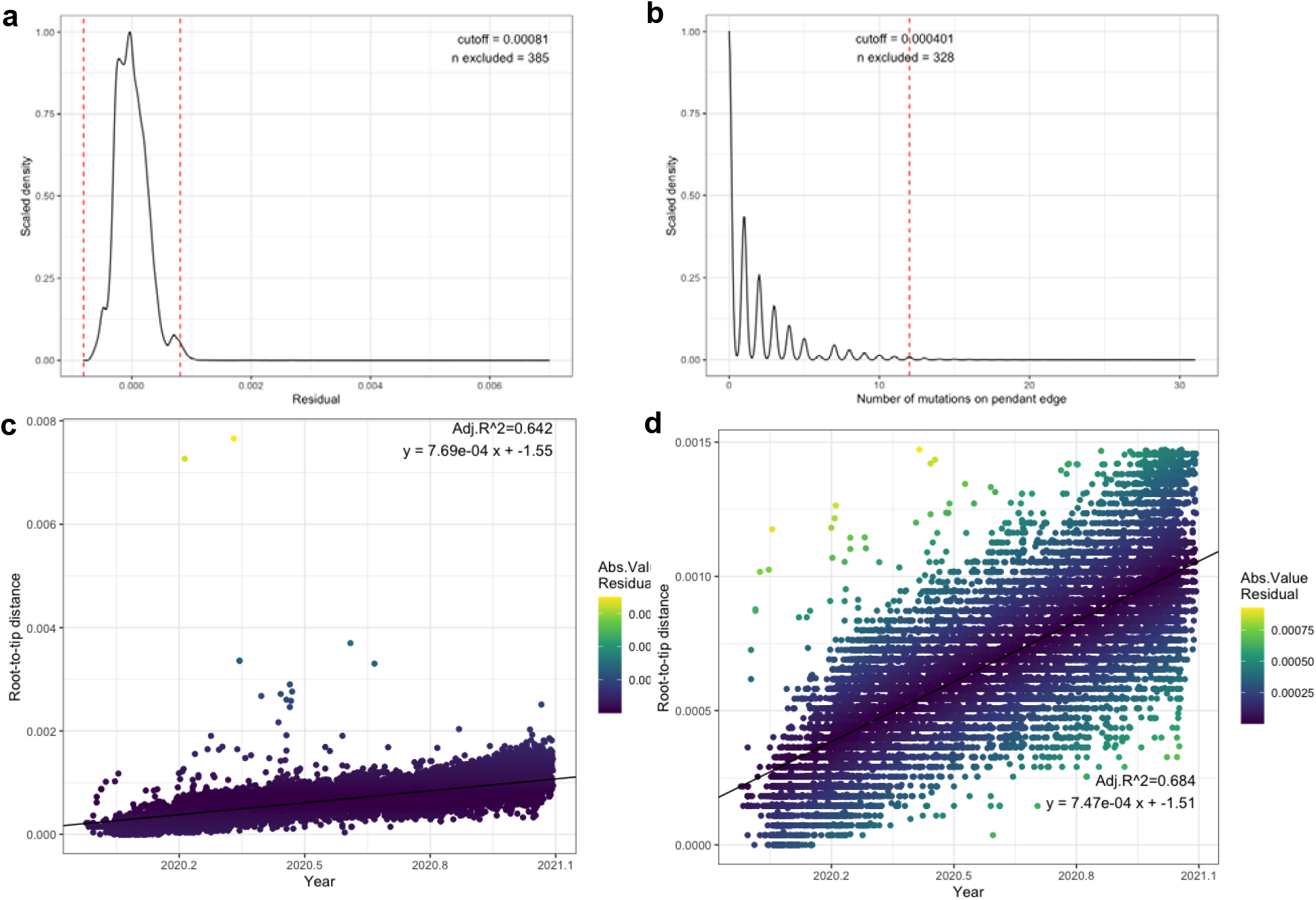
From the initially inferred FastTree phylogeny, temporal outliers (sequences that may contain excessive sequencing errors) were removed from the clock signal if a) tips residual value from the linear regression of root-to-tip distance over time exceeded the mean with three standard deviations, or if b) pendent edges (branch preceding tip) exceeded 12 mutations. The clock signal fit improved from c) the pre-trimmed tree, and d) the final tree excluding temporal outliers. The mean substitution rate was estimated to be 7.58e-4 subs/site/year.

**Fig. S5.**
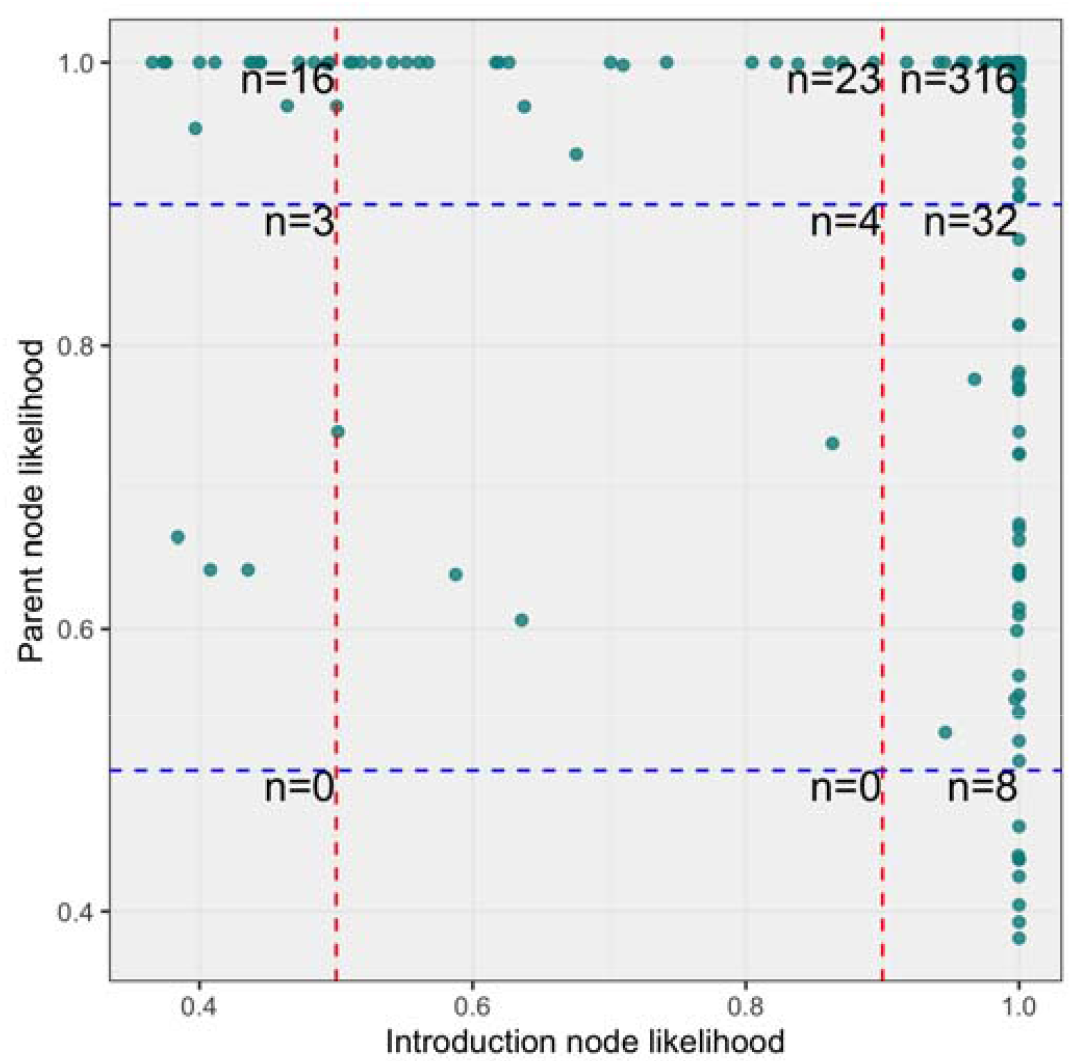
Node and parent likelihoods from a representative bootstrap of the ancestral state reconstruction.

**Fig. S6.**
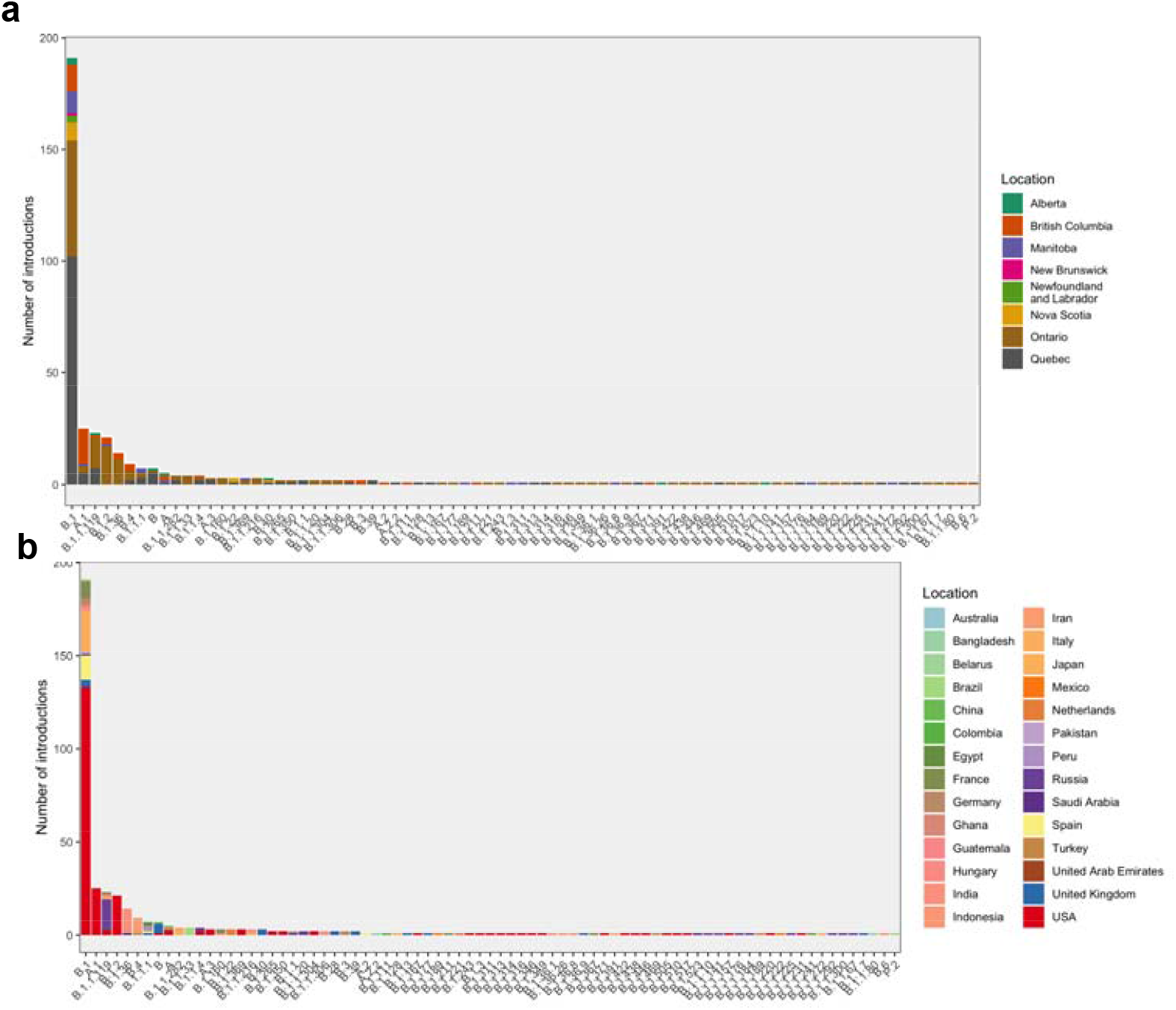
Sublineage introduction origins ordered by lineage and colored by a) province of introduction, and b) location of origin (vice versa to Fig. 3).

**Fig. S7.**
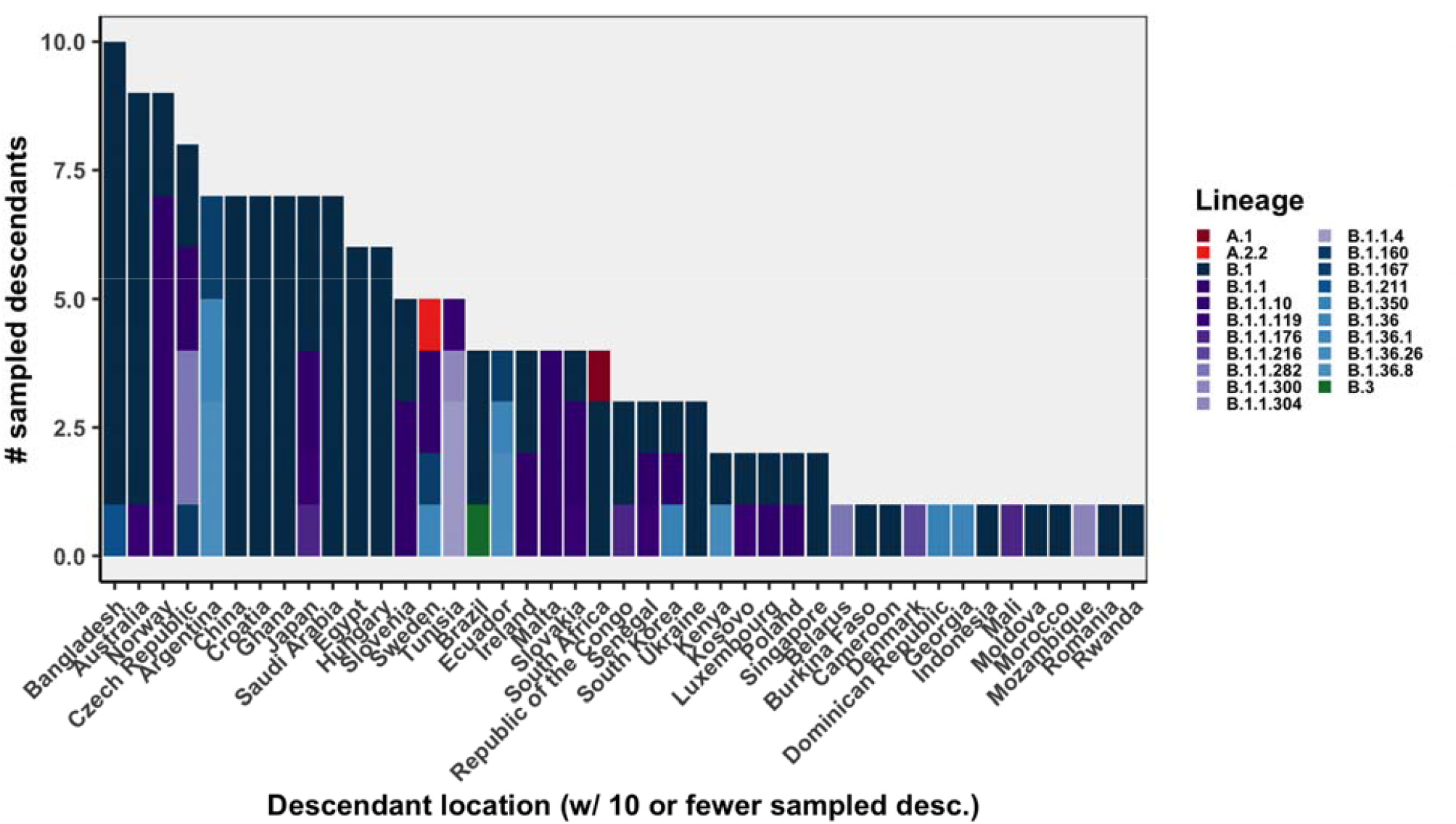
Descendant locations by lineage for locations with fewer than 10 descendants from a Canadian sublineage (only those with 10+ descendants shown in Fig. 3).

**Fig. S8.**
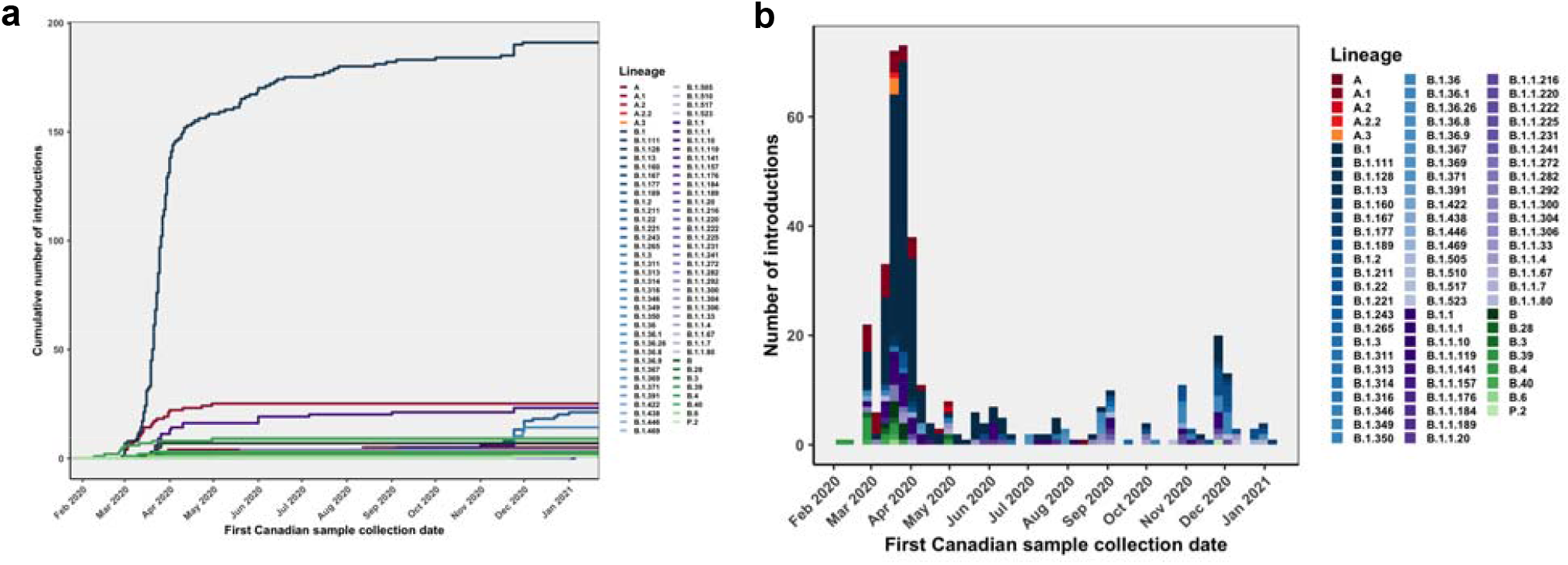
Introduction lineages identified over time, similar to Fig. 4, which shows locations over time.

**Fig. S9.**
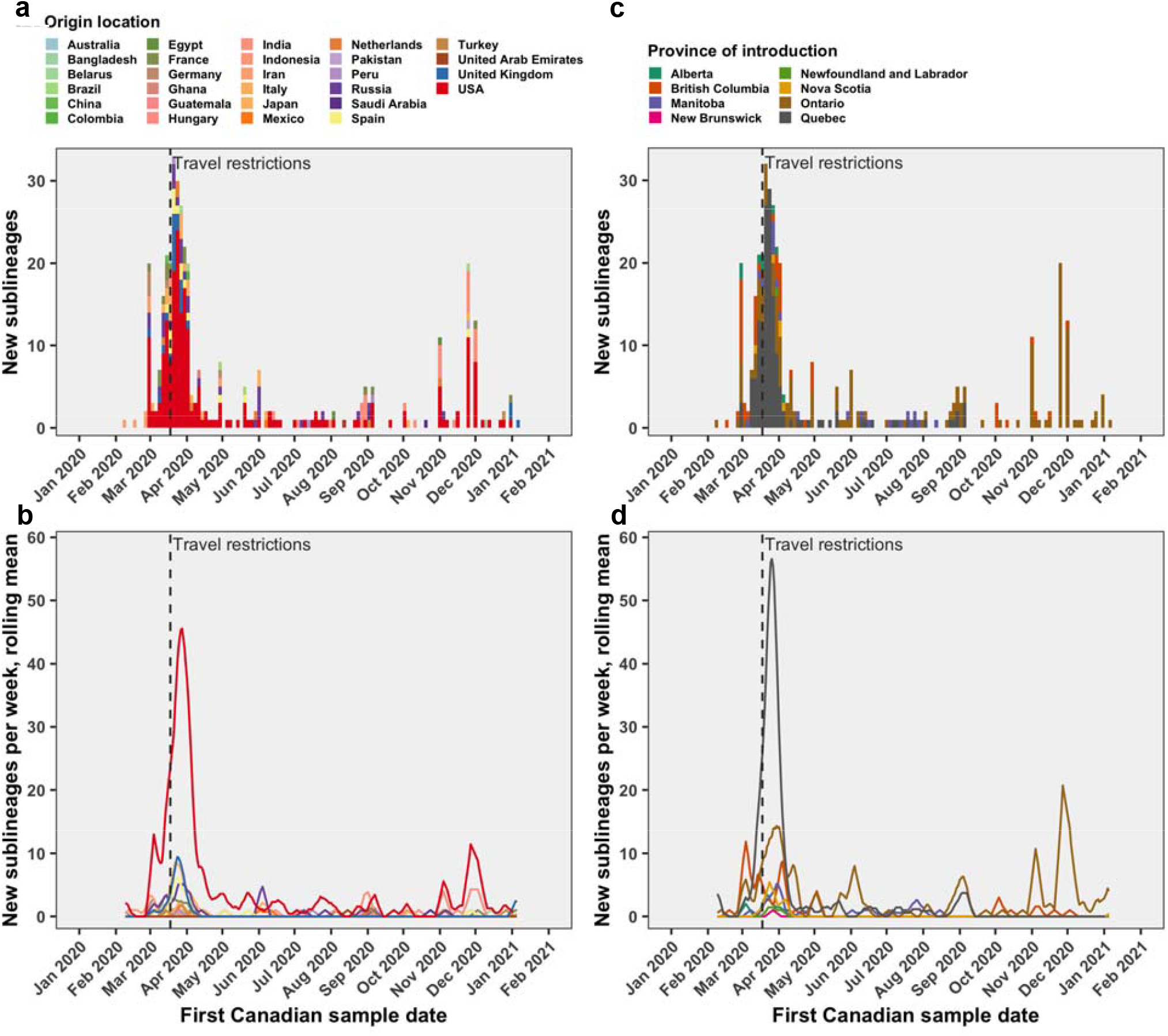
Tracking the first Canadian sample date instead of the date of the most recent common ancestor of introductions resulting in domestic transmission, aka Canadian sublineages, as in Fig. 4.

**Fig. S10.**
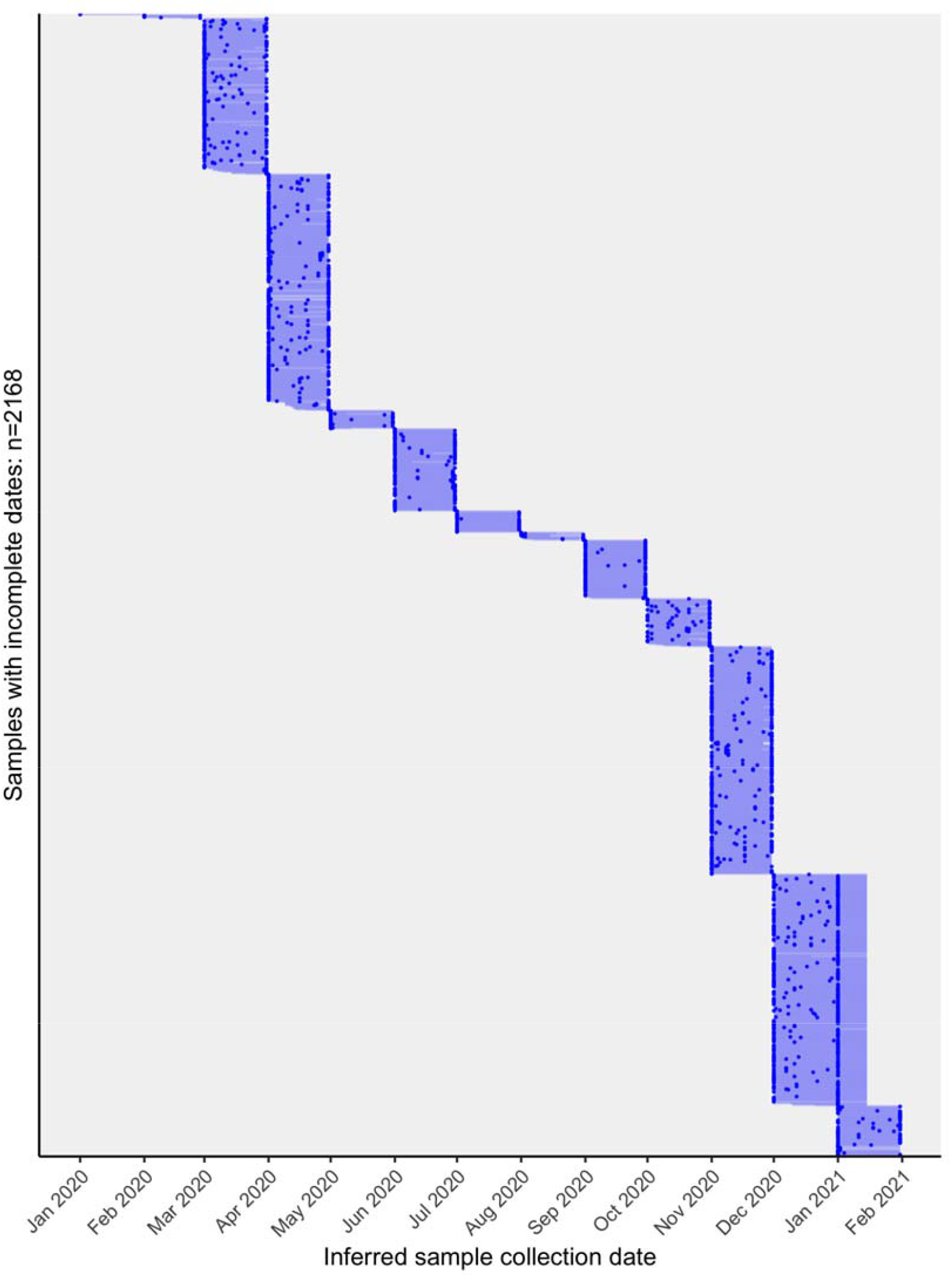
Inferred dates for 2,168 Canadian sequences that were annotated with incomplete sample collection dates containing only year and month. The blue points are the most likely LSD2-inferred date and the light blue lines show the 95% confidence interval (estimated using 100 bootstraps). The LSD-inferred dates were used to estimate the detection lag between the tMRCA and sample collection date. In instances where LSD output an estimated upper confidence date as only ‘2021’, we ascribed them arbitrarily as 15 January 2021.

**Fig. S11.**
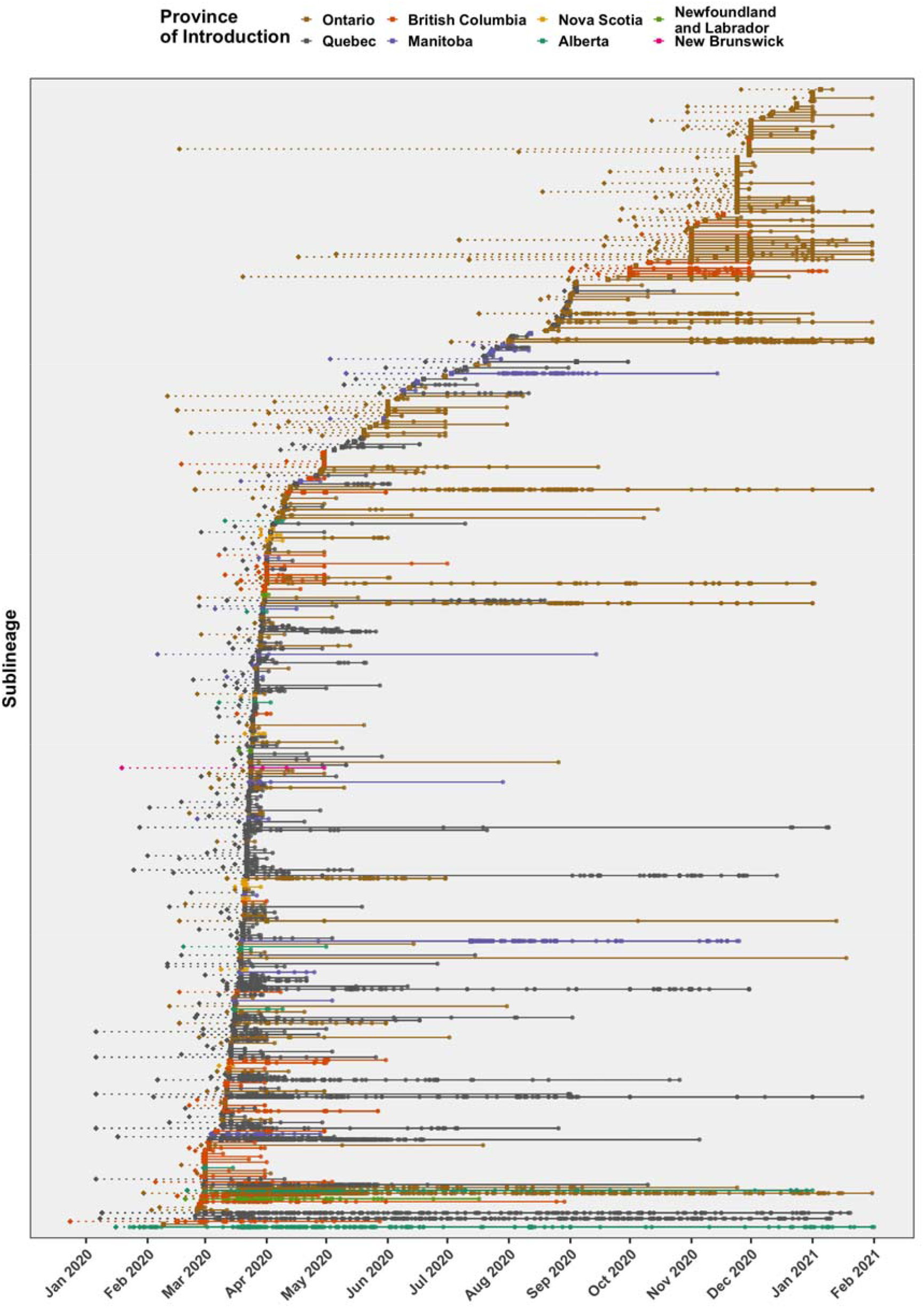
The detection lag (dotted line) was calculated as the number of days between the date of most recent common ancestor (diamond) and first Canadian sample collection date (square) for each sublineage, followed by subsequently detected descendant sample collection dates (circles), linked by solid line. The uncertainty in tMRCA and inferred sample dates was omitted for conciseness.

**Fig. S12.**
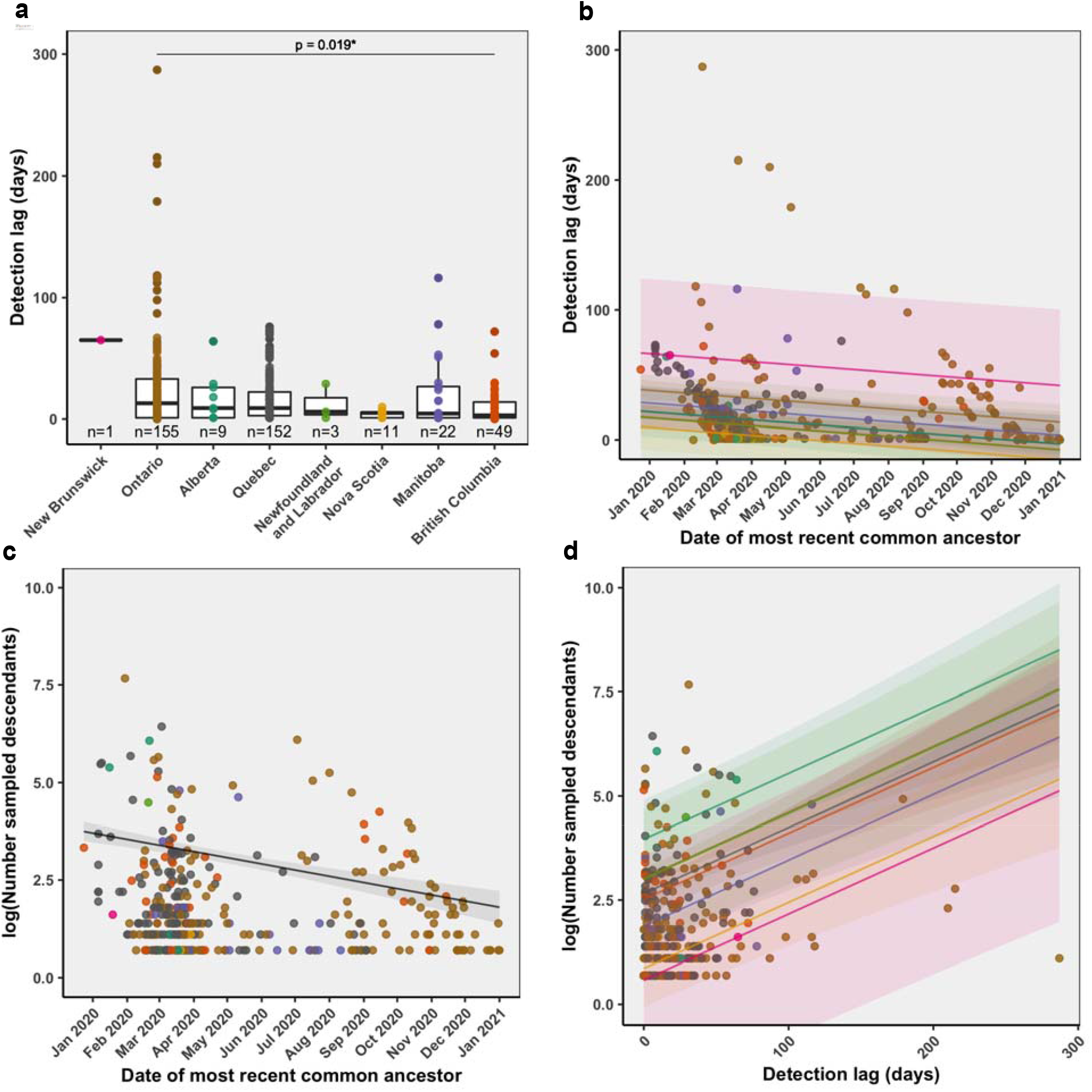
Evaluating relationships between the a) detection lag and province, b) detection lag and tMRCA, c) number of sampled descendants and tMRCA, and d) detection lag and number of sampled descendants, adjusted by province. Where detection lag was the outcome, linear models were used; where number of sampled descendants was the outcome, negative binomial models were used. The detection lag was calculated as the number of days between the first Canadian sample collection date and the date of the most recent common ancestor for each sublineage.

**Table S4.** Coefficients, 95% confidence intervals, and p-values from multiple linear regression model of sublineages’ detection lag by the tMRCA, adjusted for province of introduction (Fig. S12b). The reference category for was Ontario.

|  | Estimate | Lower 95% bound | Upper 95% bound | p-value |
| --- | --- | --- | --- | --- |
| **Intercept** | **1261.765** | **602.721** | **1920.809** | **1.93e-04** |
| **tMRCA** | **-0.067** | **-0.103** | **-0.031** | **2.6e-04** |
| **Quebec** | **-16.59** | **-23.972** | **-9.208** | **1.29e-05** |
| **British Columbia** | **-20.992** | **-30.752** | **-11.232** | **2.93e-05** |
| **Manitoba** | **-9.825** | **-23.128** | **3.477** | **1.47e-01** |
| **Nova Scotia** | **-28.497** | **-46.775** | **-10.219** | **2.33e-03** |
| **Alberta** | **-16.504** | **-36.655** | **3.647** | **1.08e-01** |
| **Newfoundland and Labrador** | **-21.166** | **-54.918** | **12.585** | **2.18e-01** |
| **New Brunswick** | **28.215** | **-29.801** | **86.231** | **3.4e-01** |

**Table S5.**
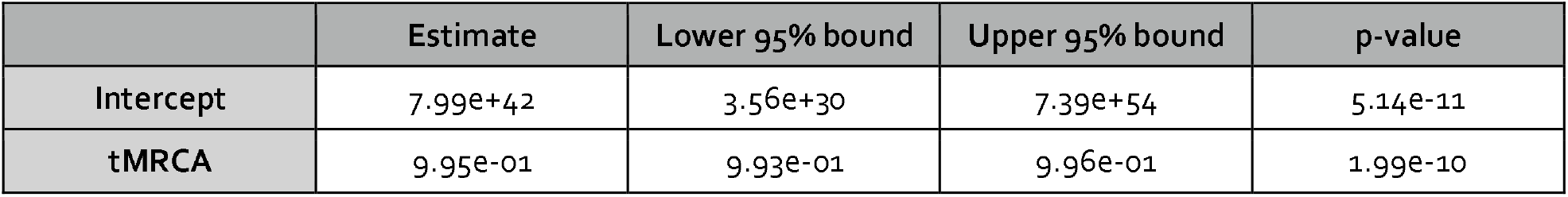
Exponentiated coefficients, 95% confidence intervals, and p-values from negative binomial model of sublineages’ number of sampled descendants by tMRCA (Fig. S12c).

**Table S6.** Exponentiated coefficients, 95% confidence intervals, and p-values from negative binomial model of sublineages’ number of sampled descendants by the detection lag, adjusted for province of introduction (Fig. S12d). The reference category for province was Alberta.

|  | Estimate | Lower 95% bound | Upper 95% bound | p-value |
| --- | --- | --- | --- | --- |
| **Intercept** | 52.449 | 22.81 | 161.973 | 3.21e-16 |
| **Detection lag** | 1.016 | 1.009 | 1.024 | 1.17e-10 |
| **British Columbia** | 0.237 | 0.073 | 0.605 | 6.26e-03 |
| **Manitoba** | 0.124 | 0.035 | 0.366 | 3.05e-04 |
| **New Brunswick** | 0.034 | 0.003 | 5.621 | 3.37e-02 |
| **Newfoundland and Labrador** | 0.383 | 0.068 | 4.103 | 3.23e-01 |
| **Nova Scotia** | 0.045 | 0.011 | 0.173 | 4.88e-06 |
| **Ontario** | 0.393 | 0.126 | 0.927 | 6.04e-02 |
| **Quebec** | 0.272 | 0.087 | 0.641 | 8.81e-03 |

**Fig. S13.**
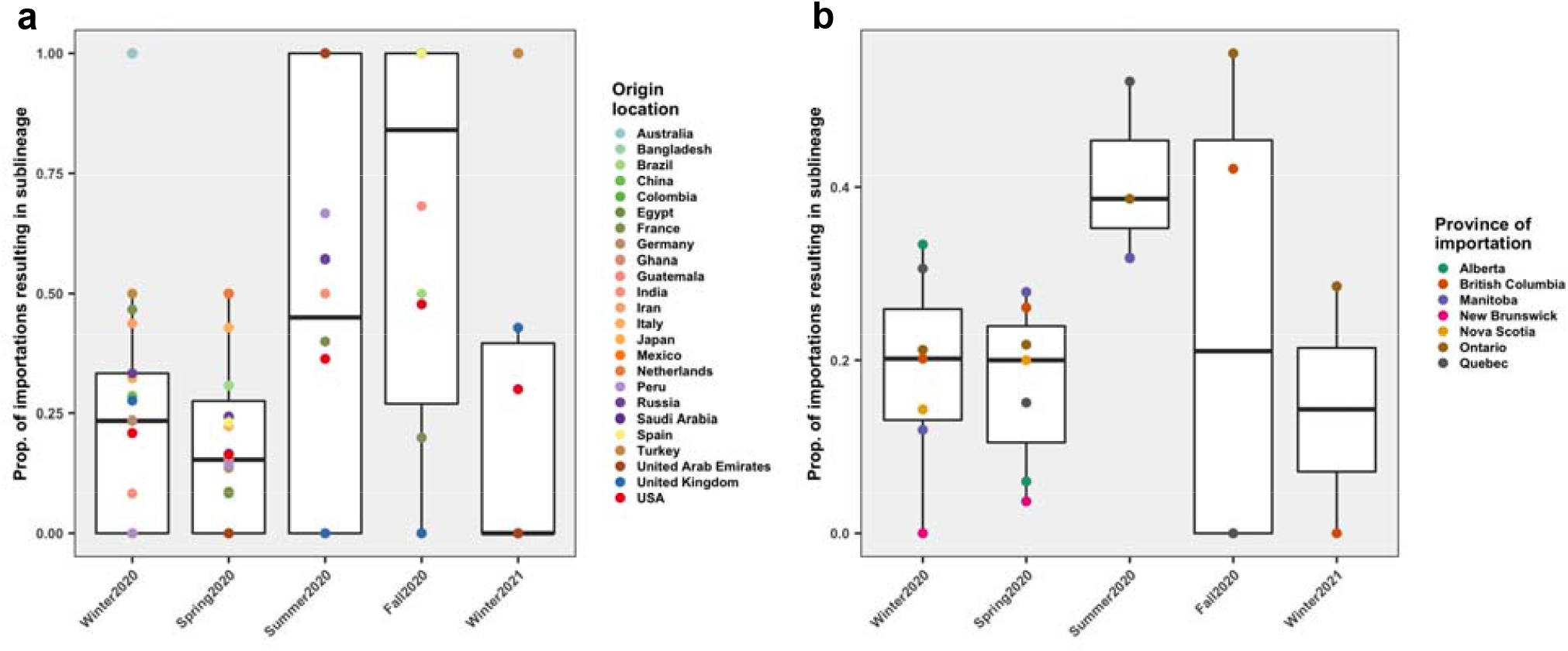
Proportion of importations resulting in sublineages (sampled domestic transmission) by a) origin location or b) province of introduction by season. Non-parametric Kruskal-Wallis tests were applied to test whether the distribution of proportion of importations resulting in singletons varied seasonally in 2020.

**Fig. S14.**
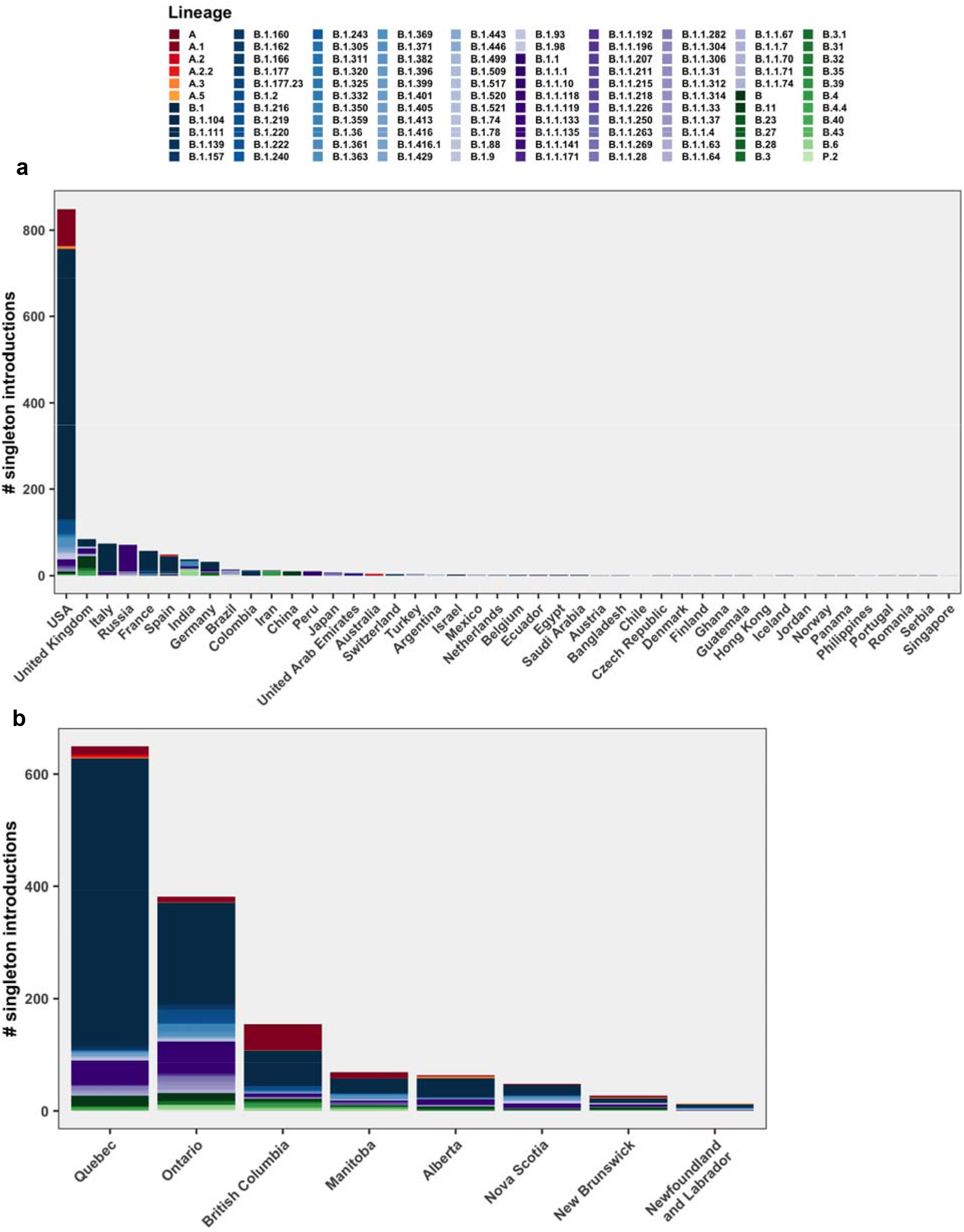
Singleton introductions by a) origin location, b) province of introduction, colored by Pango lineage.

**Fig. S15.**
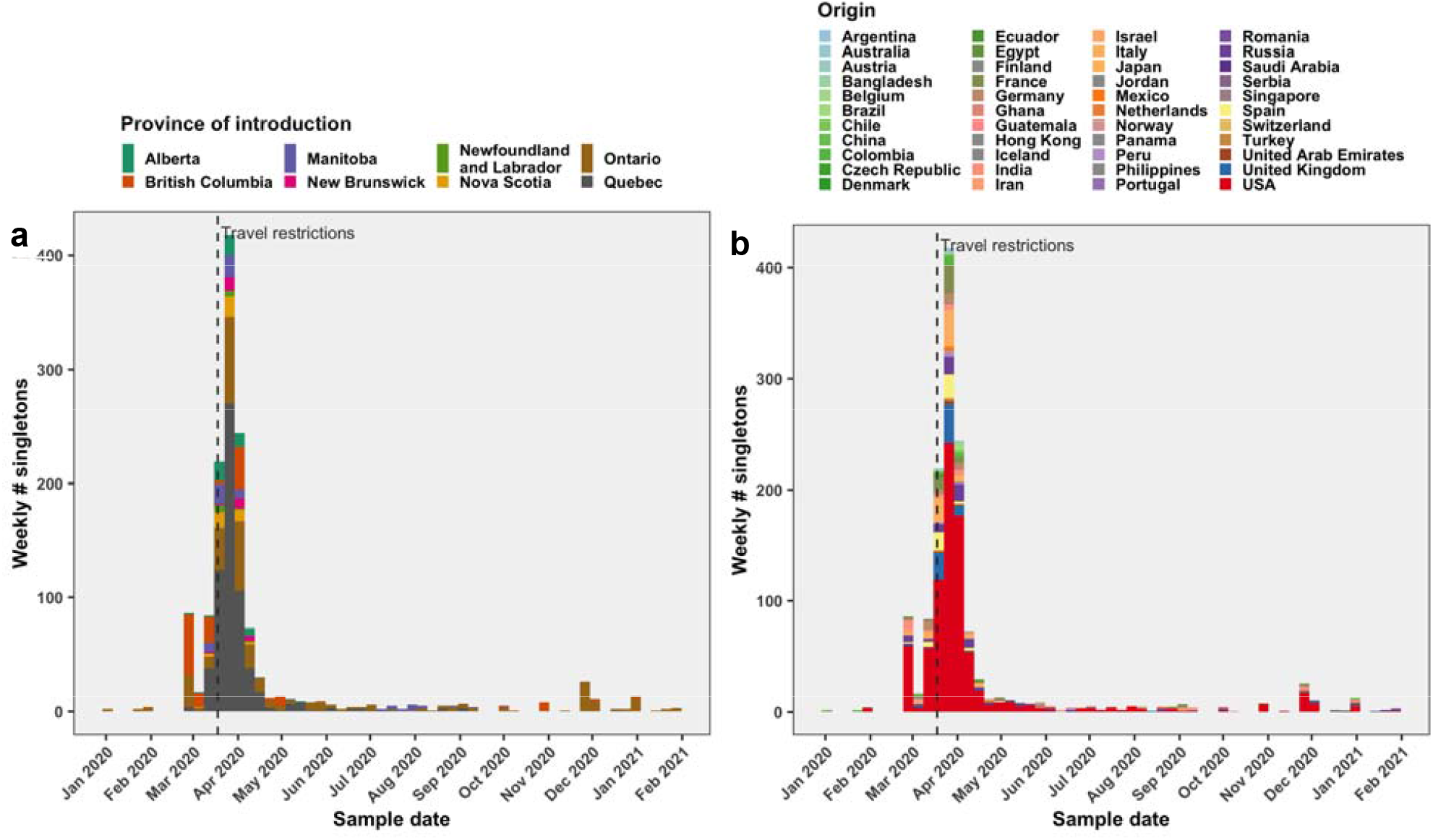
Singletons over time by a) province of introduction (tip state) and b) origin location. Canadian tips with direct international origins (as far as we sampled) that were not a descendent of a Canadian sublineage were deemed as singletons.

**Fig. S16.**
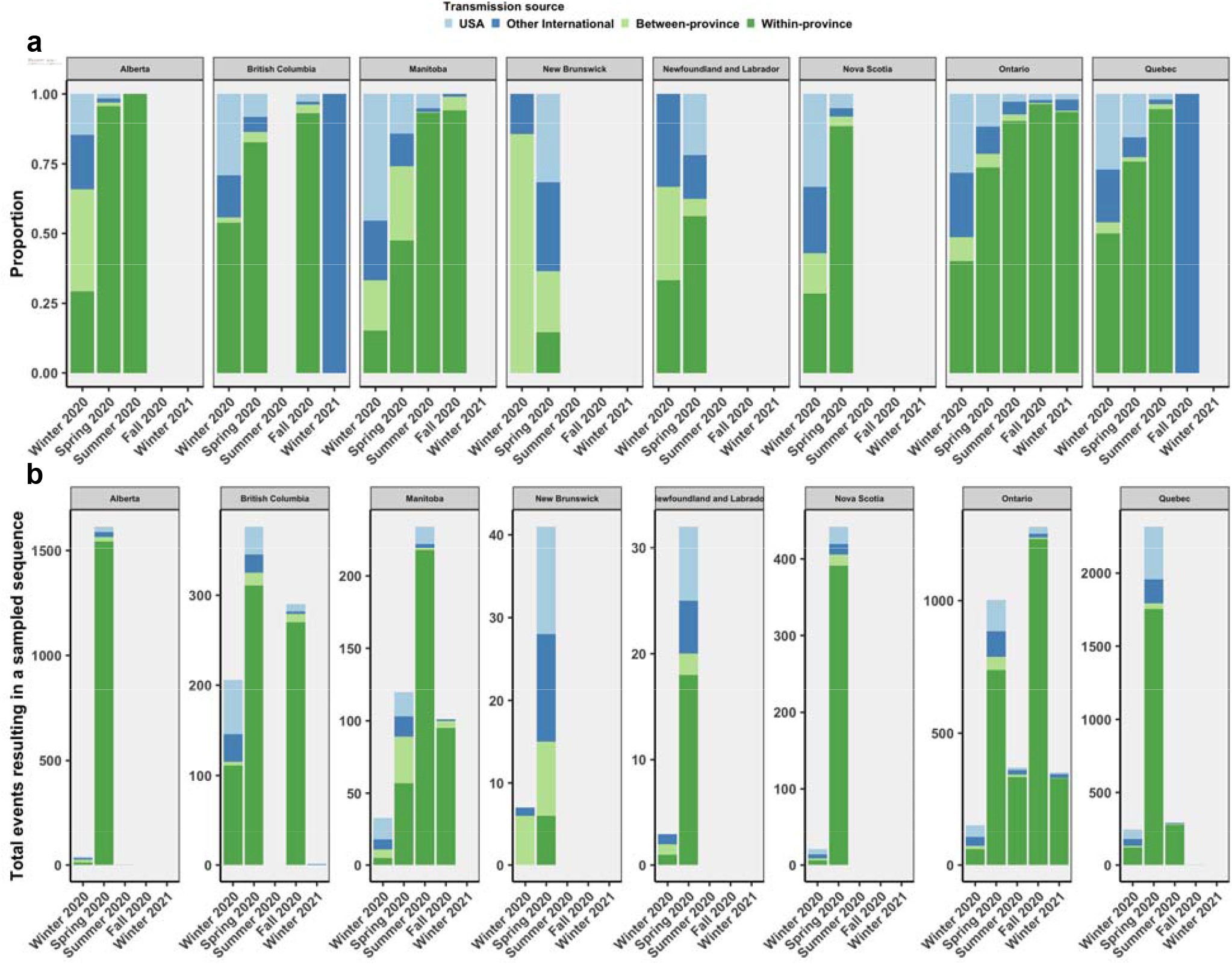
Relative contribution of transmission sources among all sampled tips by province and season. The a) proportion and b) total number of international (including the USA) transmission origins were displayed in the heatmaps in Fig. 5.

